# Rates, risks and routes to reduce vascular dementia (R4VaD), a UK-wide multicentre prospective observational cohort study of cognition after stroke: baseline data and statistical analysis plan (ISRCTN18274006)

**DOI:** 10.1101/2024.04.30.24306637

**Authors:** Philip M Bath, Ellen Backhouse, Rosalind Brown, Lisa J Woodhouse, Fergus Doubal, Terence J Quinn, Hugh S Markus, Richard McManus, John T O’Brien, Thompson Robinson, David J Werring, Nikola Sprigg, Adrian Parry-Jones, Rhian M Touyz, Steven Williams, Yee-Haur Mah, Hedley Emsley, Joanna M Wardlaw, the R4VaD Investigators

## Abstract

**Background:** Stroke is often followed by vascular cognitive impairment and vascular dementia; these are the most feared complication of stroke. However, there is limited understanding of post-stroke cognitive impairment.

**Methods:** Rates, Risks and Routes to Reduce Vascular Dementia (R4VaD) is an observational cohort study of post-stroke cognition. Patients with haemorrhagic or ischaemic stroke, or transient ischaemic attack, were recruited within six weeks of stroke from hospitals across the UK. Consent was obtained from patients with capacity or from relatives/friends in those without capacity. The primary outcome is cognition and its severity assessed using a 7-level ordinal outcome. Final cognition will be compared in those with mild stroke/TIA (worst NIHSS <=7) versus severe stroke (NIHSS >7). Secondary outcomes will include function, mood and quality of life.

**Results:** We recruited 2441 patients from 50 hospitals. Of these, 2437 (99.8%) had a qualifying event of stroke or TIA. The mean age was 68.2 years (standard deviation 13.5), females 981 (40.3%), onset to recruitment 6 days [interquartile range 3-13] and diagnosis ICH 193 (7.9%), ischaemic stroke 2101 (86.2%), TIA 143 (5.9%). The distribution of cognition at baseline was: normal 1256 (51.6%), minor neurocognitive disorder-single domain 530 (21.8%), minor neurocognitive disorder-multi domain 320 (13.1%), major neurocognitive disorder-mild 237 (9.7%), major neurocognitive disorder-moderate 90 (3.7%) and major neurocognitive disorder-severe 3 (0.1%). We provide the statistical analysis plan in the appendix.

**Conclusion:** We provide baseline data and the SAP. Final follow-up will be completed in quarter 2 2024. The data highlight the substantial under-appreciated cognitive burden of stroke, even in the first few days and weeks.

## INTRODUCTION

Stroke, transient ischaemic attack (TIA), post-stroke cognitive impairment (PSCI) and post-stroke dementia (PSD) share many risk factors,^1^ and are risk factors for each other.^2 3^ Nevertheless, risk-prediction for the development of PSCI and PSD after stroke/TIA remains challenging.^2 4^

Although cognitive impairment is the top concern of many stroke survivors,^5^ data are limited regarding rates and progression of PSCI by time and clinically-relevant strata that account for pre-morbid and pre-stroke cognition, medical, lifestyle and socioeconomic factors.^2^ Improved understanding of these predictors would improve risk stratification and identify mechanisms and intervention targets.^1^

The limited progress to date in predicting, understanding and ameliorating PSCI reflects the variability of stroke presentations, its consequences and co-morbidities. Nonetheless, PSCI is a common, under-researched problem and a priority for patients, carers, health services, funders, policy makers and governments in recent years.^6^

The Rates, Risks and Routes to Reduce Vascular Dementia (R4VaD) study is a Priority Programme in VaD funded by the Stroke Association, British Heart Foundation and Alzheimer’s Society. It is a large, multicentre, longitudinal, representative study in patients presenting with recent stroke or TIA to UK Stroke Centres and aiming to assess cognition, functional and neuropsychiatric outcomes for up to two years after stroke using standardised proportionate ascertainment methods.^7^

Prior to presentation of the primary results, this publication presents: i) a detailed listing of baseline characteristics; and ii) the statistical analysis plan (SAP) in the accompanying Supporting Information Appendix S1. The SAP is presented prior to locking of the study database so that analyses are not data driven or reported selectively.^8^ In addition to the SAP, we also list planned secondary analyses and substudies.

## METHODS

The protocol is published in detail ^7^ so we only provide brief details on study aims, design and approvals, eligibility, consent, baseline characteristics and their analysis here.

### Study aims

The primary aim of R4VaD was to determine rates of cognitive impairment and dementia up to at least one year after stroke and, in most participants, two years. Secondary aims included identifying key risk predictors and mechanisms for the development of PSCI and VaD, improve cognitive testing for PSCI, provide data to inform the design of future randomised controlled trials and clinical services for patients with PSCI, and establish a well phenotyped population with consent for re-contact for future studies and trials.

### Design

R4VAD is a large multicentre longitudinal study in patients presenting with stroke or TIA to UK stroke centres. The study has ethics approval in England, Northern Ireland, Scotland and Wales, NHS Research and Innovation Office approvals in each participating site and was adopted by the National Institute for Health and care Research (NIHR) Clinical Research Network, or equivalent across the UK. All participants or their appropriate guardian, and informants, give written informed consent.

### Eligibility criteria

Adult in-patients assessed at UK stroke centres with ischaemic stroke, spontaneous intracerebral haemorrhage (ICH) or TIA, who were expected to survive to at least 12 weeks after stroke, and irrespective of cognitive function and ability to consent, were eligible. Patients with stroke mimicking conditions, subarachnoid haemorrhage or secondary brain haemorrhage were excluded. Co-enrolment into a randomised controlled trial and other observational studies was allowed.

### Consent

R4VaD was approved by Ethics Committees in Scotland (A Research Ethics Committee; Ref 18/SS/055), England (Health Research Authority), Wales (Health and Care Research) and Northern Ireland (all Northeast Newcastle and North Tyneside 1; 18/NE/0150). NHS Research and Innovation Office approval was given in each participating site. R4VaD is registered (ISRCTN18274006).

Participants with capacity (and informants where available) gave written informed consent at study entry. Proxy consent for patients who were deemed to lack capacity was obtained from family, friends or legal guardian according to national rules. The consentee could also act as the informant.

### Baseline characteristics

Demographic (including age, sex, ethnicity), medical history (vascular risk factors, prior stroke/TIA, prior cognitive impairment/dementia, blood pressure), lifestyle, stroke-related findings (subtype, severity [National Institutes of Health stroke scale, NIHSS]), education (highest level), cognitive (impairment, dementia, Montreal cognitive assessment [MoCA], verbal fluency, Trails A and B, telephone interview cognitive scale-modified [TICS-M], 4- and 7-level cognition scale [Supplementary Figure 1] ^7 9^), functional measures (modified Rankin scale [mRS], clinical frailty scale [CFS]) and imaging (type, acute lesion features, pre-stroke features [atrophy, white matter hyperintensities [WMH], prior stroke) and other investigation (carotid imaging, ECG) findings at baseline were collected into a centralised online database maintained at the Stroke Trials Unit, University of Nottingham.

### Statistical analyses

Participants who are not deemed to have entered the study with a cerebrovascular event (ICH, IS, TIA) are excluded from all analyses. Most analyses are descriptive with data shown as number (%), median [interquartile range, IQR] or mean (standard deviation, SD). The 4- and 7-level cognition scale were derived from MoCA, TICS-M, mRS, residence, clinical diagnoses during follow-up and other available measures.^7^ Comparisons of baseline characteristics by stroke severity and 7-level cognition scale were assessed using Chi-square test, Mann-Whitney U test, t-test, Jonckheere-Terpstra (JT), Kendall’s Tau and ANOVA for trend. Analyses were performed using SAS version 9.4 (SAS Institute, Cary, NC, USA).

## RESULTS

The study commenced recruitment in October 2018 and completed recruitment in September 2022. There was a temporary hold to recruitment in March 2020 due to COVID-19; the subsequent recruitment rate was lower than pre-COVID (Figure 1). Final follow-up is planned for quarter 1 2024.

**FIGURE 1.**
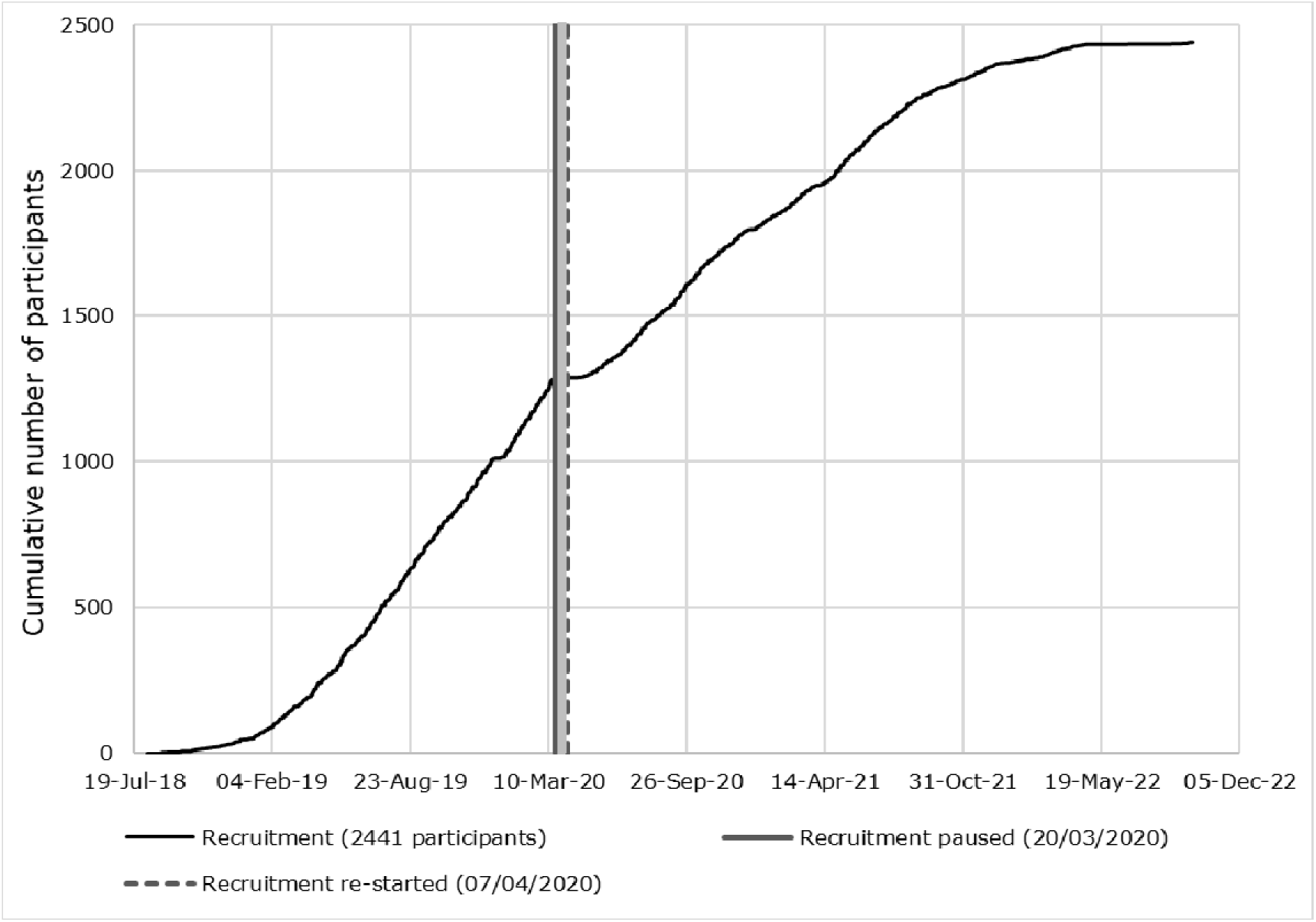
Recruitment curve.

Of a planned 2000 participants, we recruited 2441 (122%) from 50 UK hospitals. Four (0.2%) of these did not have a qualifying diagnosis stroke or TIA so were excluded from subsequent analyses. The 2437 participants with a stroke or TIA were recruited at a median of 6 [3, 13] days from onset (Table 1). At baseline, the mean age was 68.2 (13.5, range 19-97) years and there were 981 (40.3%) female patients. Vascular risk factor rates included hypertension 1543 (63.4%), hyperlipidaemia 1122 (46.1%), diabetes mellitus 544 (22.3%), atrial fibrillation 485 (19.9%), history of previous stroke 383 (15.7%) and past or current smoking 1421 (58.9%). A third (742, 31.3%) had a close family history of stroke and 378 (16.0%) a close family history of dementia. Most participants (2307, 94.7%) had capacity, and 831 (35.5%) participants had A-level equivalent school exams or higher qualifications. Baseline clinical findings included motor weakness in 1808 (74.2%, left 1050, right 699, bilateral 59), sensory change in 966 (39.6%), neglect/inattention in 453 (18.6%), dysphasia in 571 (23.4%) and visual disturbance in 450 (18.5%). The mean blood pressure was 135.3 (19.9)/76.9 (12.1) mmHg (Table 1).

**Table 1.**
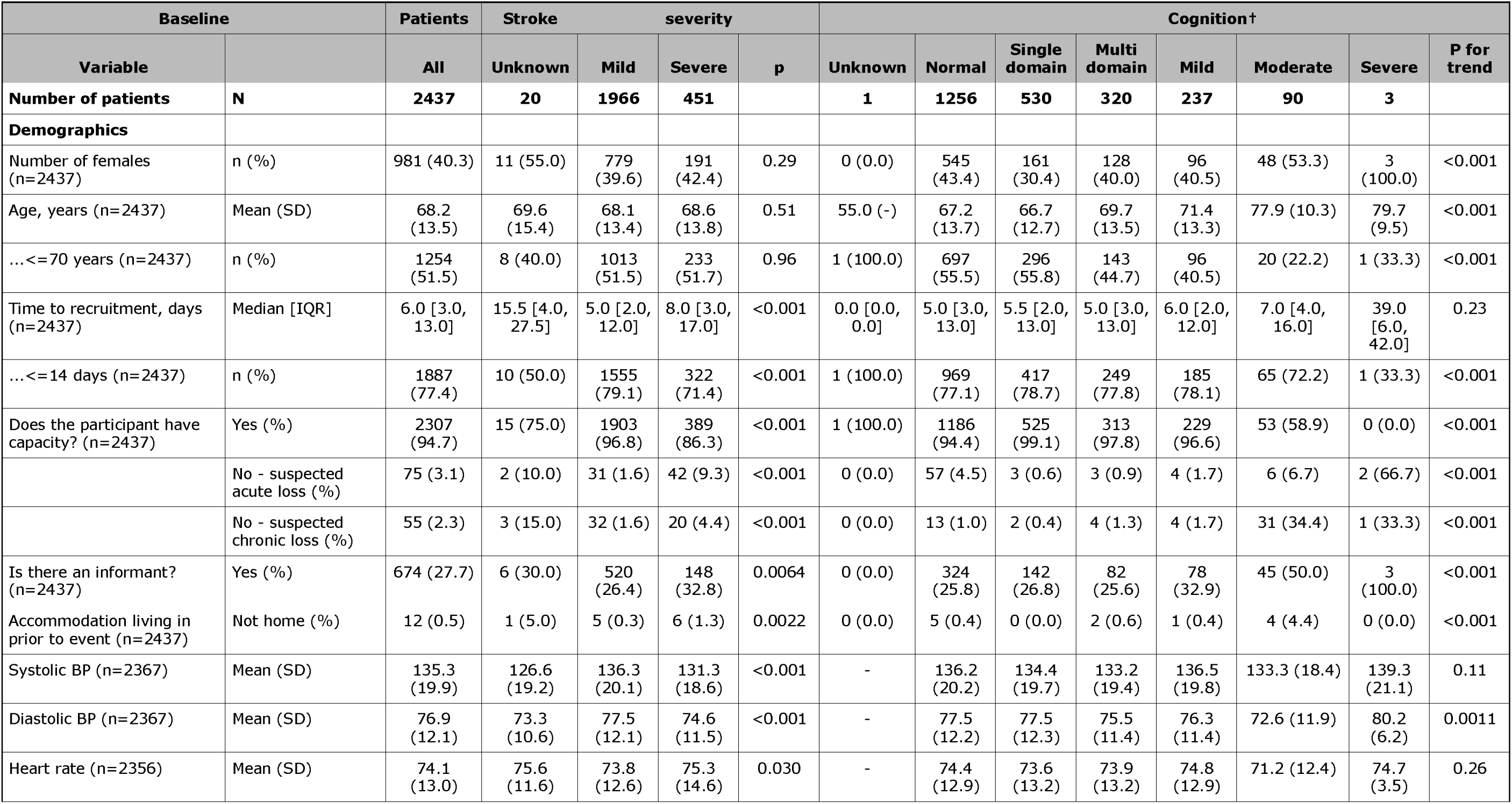
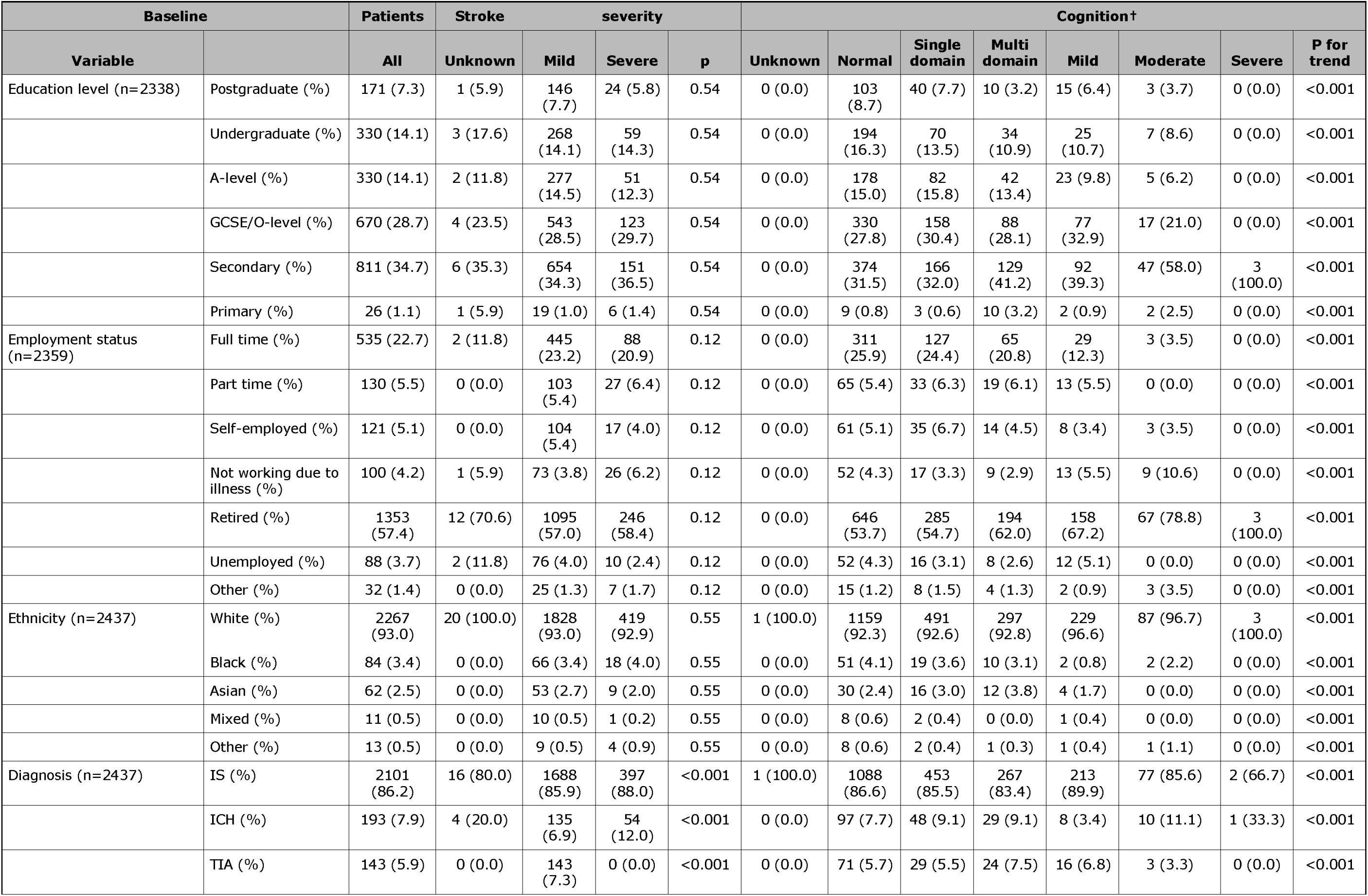
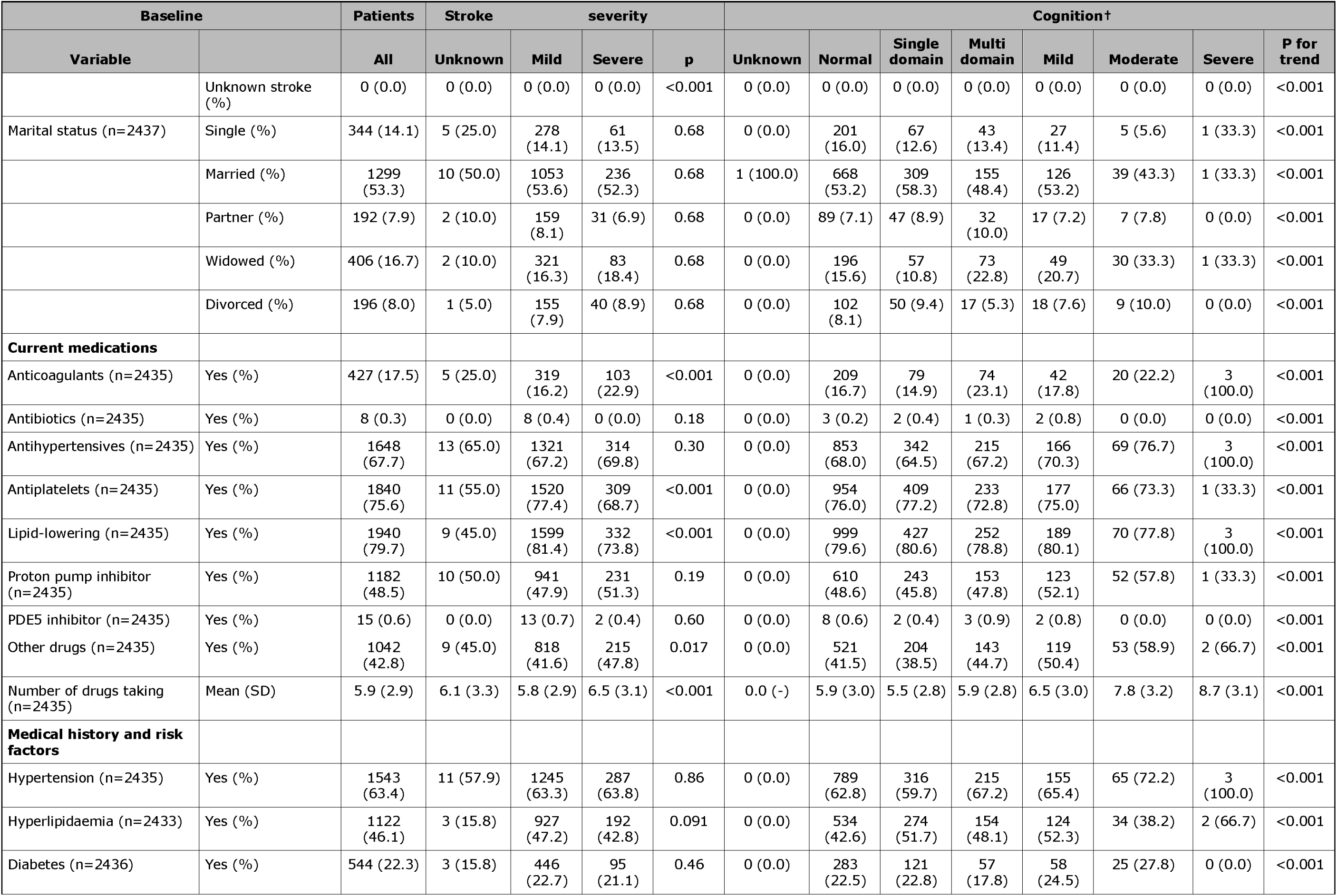
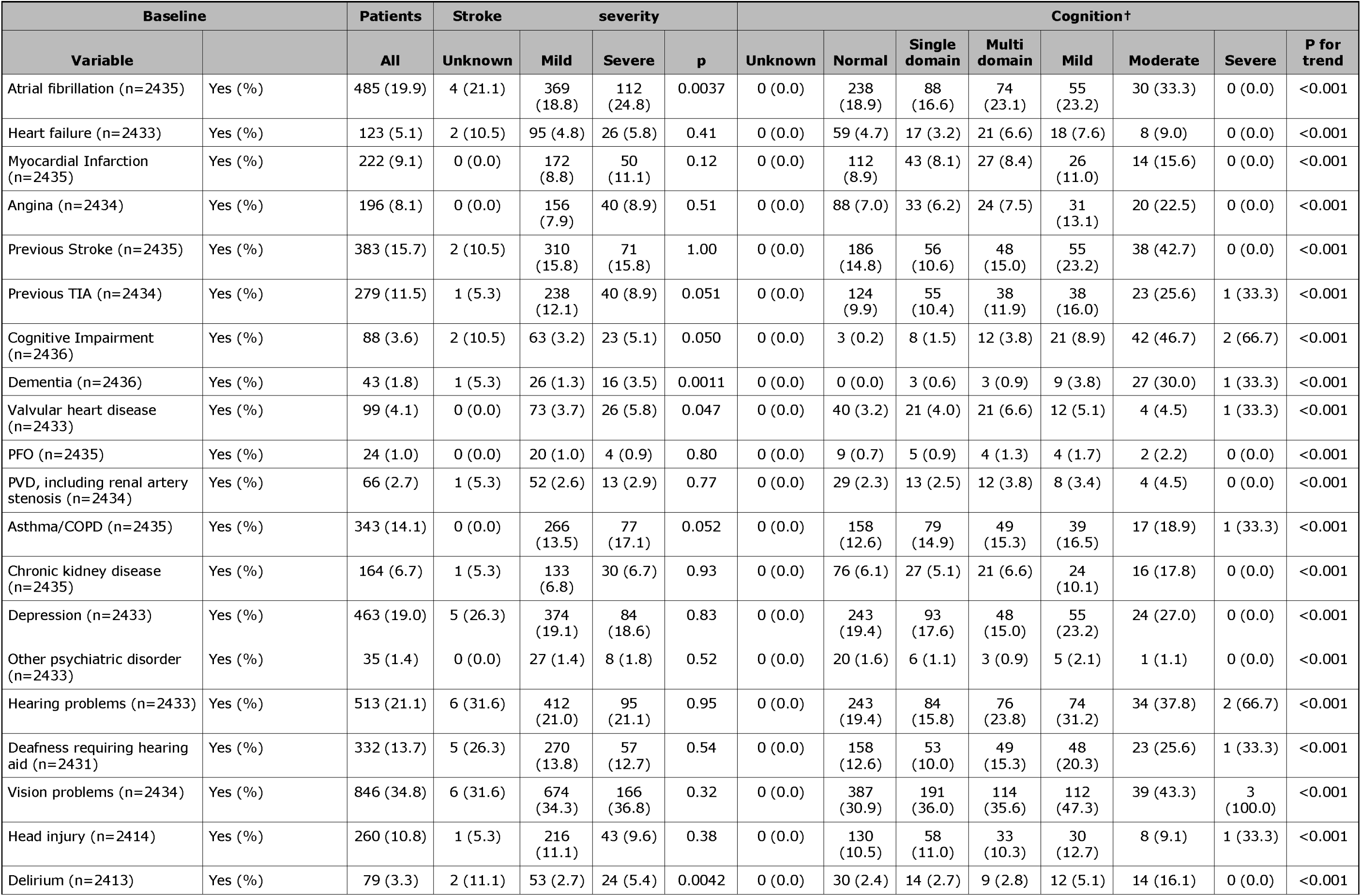
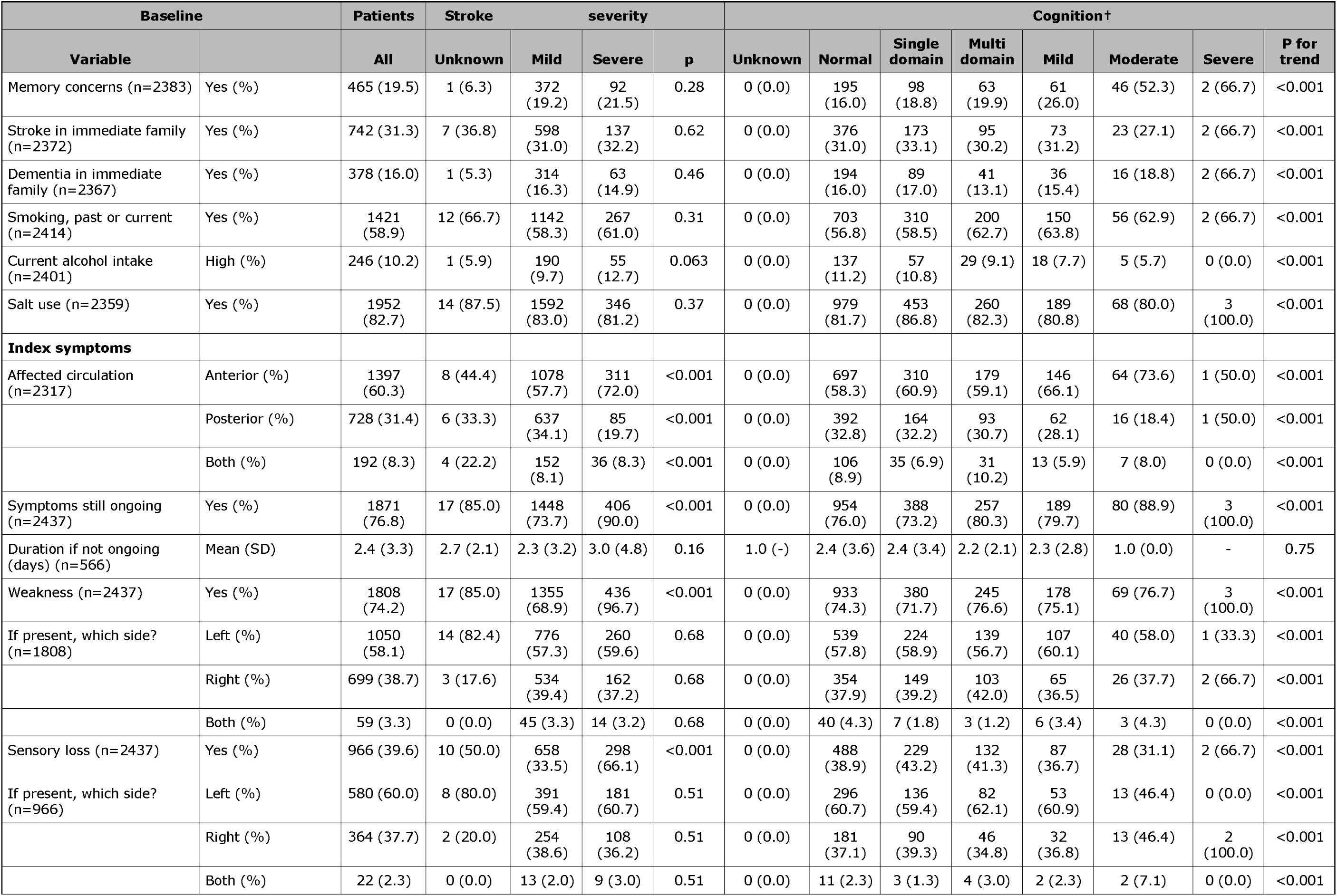
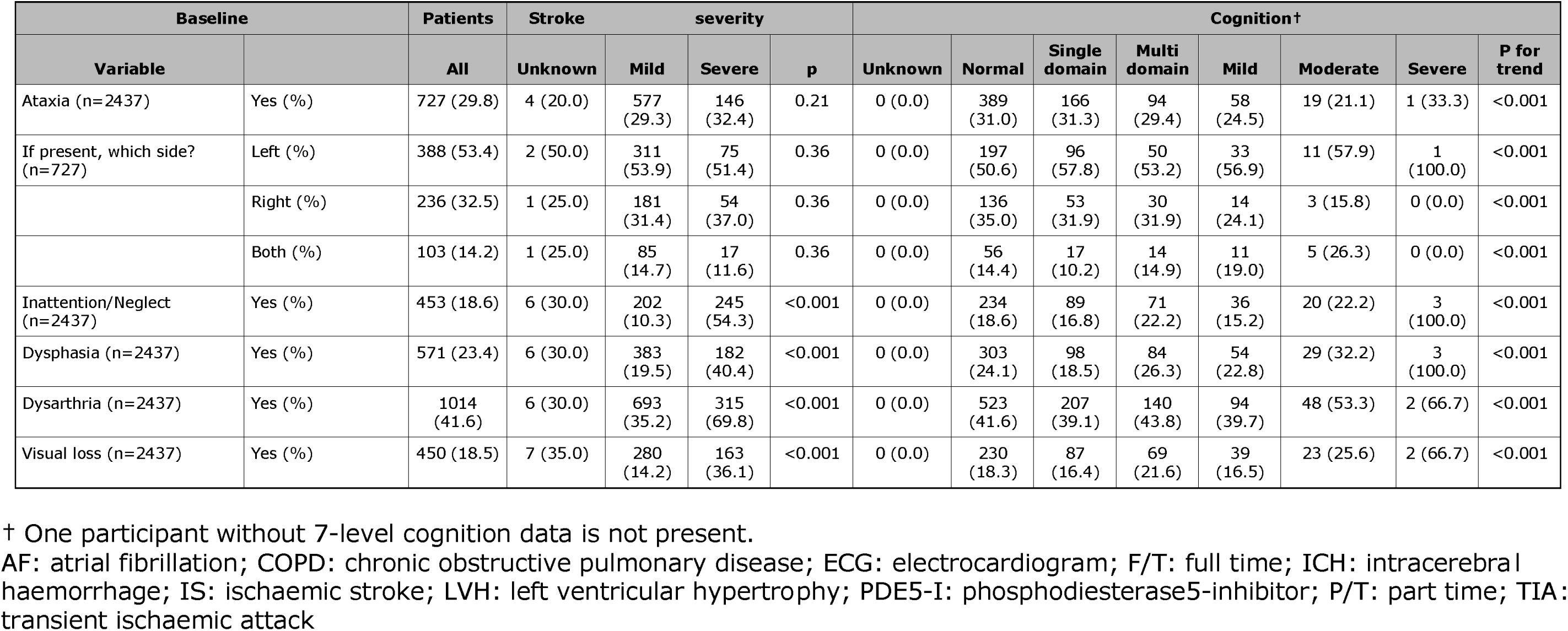
Baseline demographic characteristics in stroke or TIA by baseline stroke severity (NIHSS≤7 vs >7) and cognitive status. Data are number (%), median [interquartile range] or mean (standard deviation). Comparison by Chi-square, Mann-Whitney U test and t-test or Jonckheere-Terpstra (JT), Kendall’s Tau and ANOVA for trend.

At baseline, cognitive impairment and dementia were recorded in the medical history in 88 (3.6%) and 43 (1.8%) of participants, respectively (Table 1). On cognitive testing, the median Montreal Cognitive Assessment (MoCA) score was 24.1 (4.3) with 693 (40.6%) having a score <25 signifying cognitive impairment (Table 2). The TICS-M was 23.9 (5.1) and median score on the trail making test, part B was 20.8 (7.4) points with this achieved in 152.7 (102.6) seconds. The primary 7-level cognitive outcome distribution was: normal 1256 (51.6%), single domain minor neurocognitive disorder 530 (21.8%), multi-domain minor neurocognitive disorder 320 (13.1%), minor dementia 237 (9.7%), moderate dementia 90 (3.7%) and severe dementia 3 (0.1%) (Figure 2a). The median premorbid modified Rankin scale (mRS) was 0 [0-1] (Figure 2b). The mean worst NIHSS was 4.9 (SD 4.7, range 0-35) and 417 (19.5%) had a worst NIHSS>7 (Table 2, Figure 2c).

**FIGURE 2.**
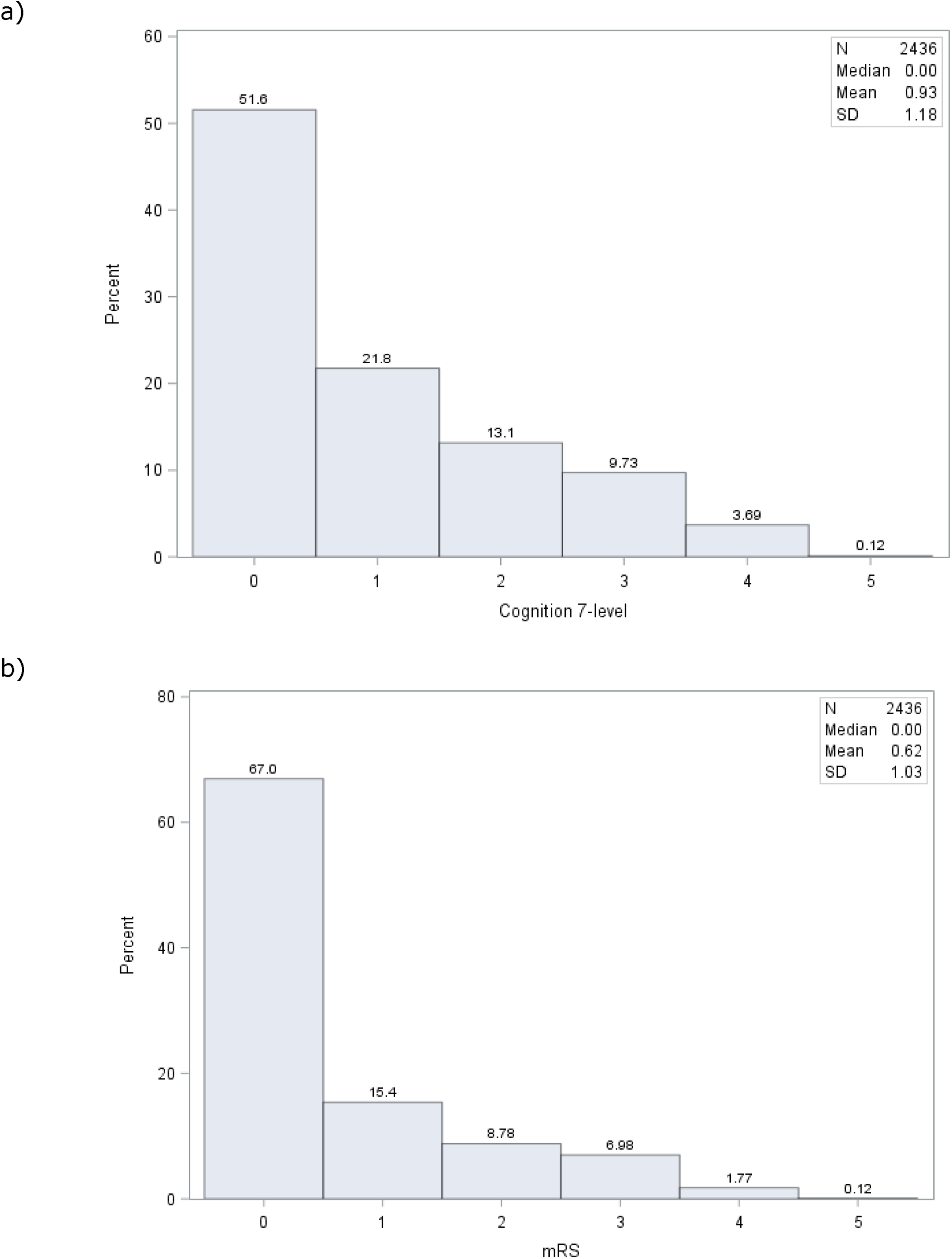
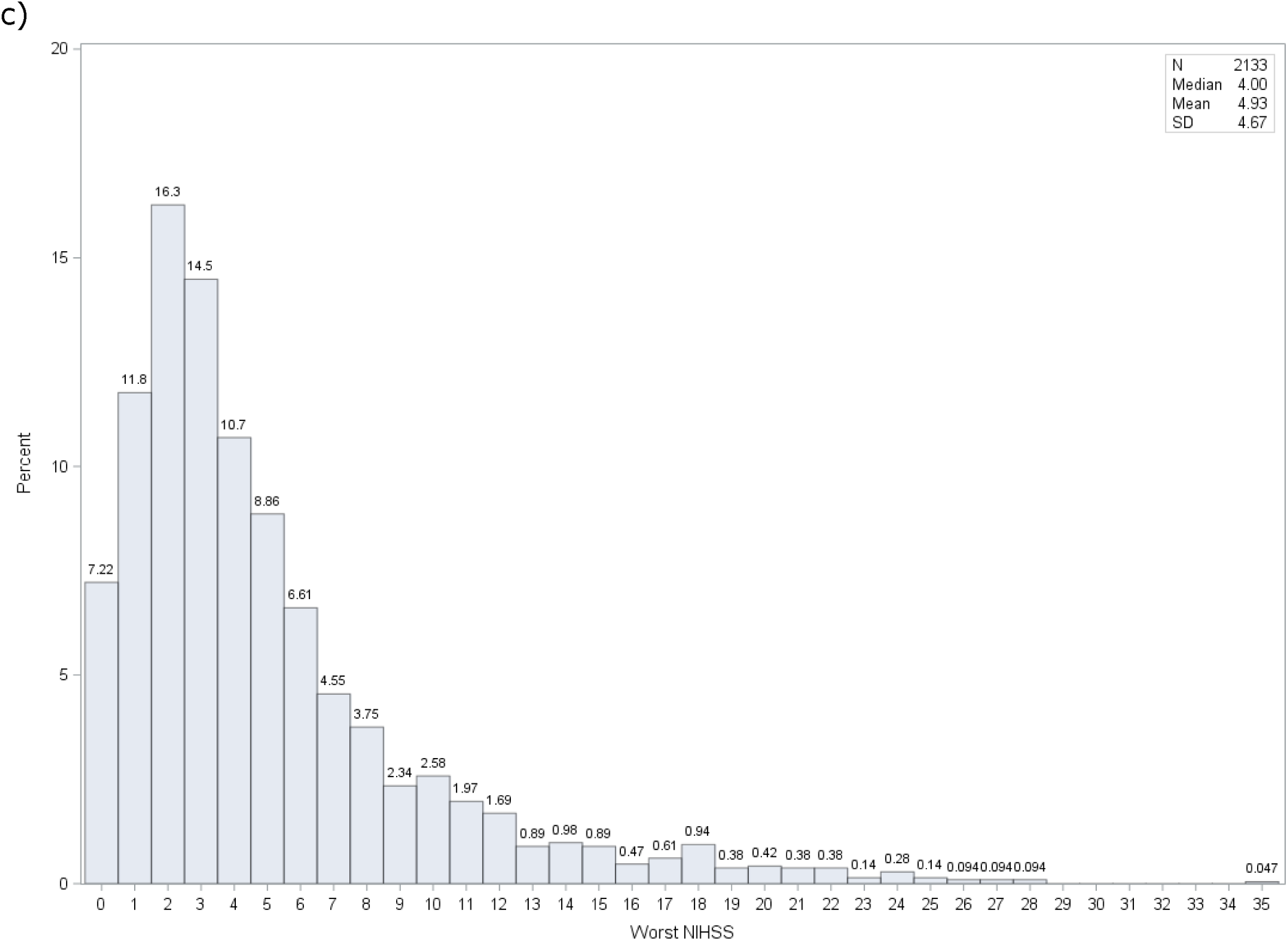
Distribution of: a) baseline 7-level cognition/dementia scale; b) pre-stroke modified Rankin Scale; b) worst post-stroke National Institutes of Health stroke scale; and c).

**Table 2.** Baseline cognitive, functional, mood and other assessments by baseline stroke severity (NIHSS≤7 vs >7) and cognitive status. Data are number (%), median [interquartile range] or mean (standard deviation). Comparison by Chi-square, Mann-Whitney U test and t-test or Jonckheere-Terpstra (JT), Kendall’s Tau and ANOVA for trend, with adjustment for age, sex, educational attainment, pre-stroke mRS, stroke type (TIA, IS, ICH), stroke severity (NIHSS), 7-level cognition and time from stroke onset to consent/assessment.

| Baseline |  | Patients | Stroke | Severity (NIHSS) |  |  | Cognition | Cognition |  |  |  |  |  |  |
| --- | --- | --- | --- | --- | --- | --- | --- | --- | --- | --- | --- | --- | --- | --- |
| Variable |  | All | Severity Unknown | Mild | Severe | p | Severity Unknown | Normal | Single domain | Multi domain | Mild | Moderate | Severe | P for trend |
| Number of patients | N | 2437 | 20 | 1966 | 451 |  | 1 | 1256 | 530 | 320 | 237 | 90 | 3 |  |
| Pre stroke |  |  |  |  |  |  |  |  |  |  |  |  |  |  |
| Quality of life |  |  |  |  |  |  |  |  |  |  |  |  |  |  |
| Pre-morbid EQ-5D-5L HSUV (n=1234) | Mean (SD) | 0.9 (0.2) | 0.8 (0.1) | 0.9 (0.2) | 0.9 (0.2) | 0.26 | - | 0.9 (0.2) | 0.9 (0.1) | 0.9 (0.1) | 0.9 (0.2) | 0.6 (0.3) | 0.7 (0.4) | <0.001 |
| Activities of daily living |  |  |  |  |  |  |  |  |  |  |  |  |  |  |
| Barthel Index (n=1024) | Mean (SD) | 94.6 (12.7) | 88.8 (24.7) | 95.9 (9.9) | 89.0 (20.2) | <0.001 | - | 95.3 (10.8) | 97.6 (8.0) | 94.1 (14.1) | 94.4 (11.8) | 71.0 (24.8) | 80.0 (-) | <0.001 |
| Lawton's ADL (n=1012) | Mean (SD) | 10.6 (4.9) | 12.8 (7.2) | 10.2 (4.5) | 12.0 (6.3) | <0.001 | - | 10.2 (4.2) | 9.1 (2.8) | 10.2 (4.5) | 11.4 (5.2) | 21.4 (7.9) | 20.0 (-) | <0.001 |
| Brief physical activity assessment, Vigorous (n=2310) | Median [IQR] | 0.0 [0.0, 2.0] | 0.0 [0.0, 0.0] | 0.0 [0.0, 2.0] | 0.0 [0.0, 2.0] | 0.84 | - | 0.0 [0.0, 2.0] | 0.0 [0.0, 2.0] | 0.0 [0.0, 2.0] | 0.0 [0.0, 2.0] | 0.0 [0.0, 0.0] | 2.0 [0.0, 4.0] | <0.001 |
| Brief physical activity assessment, Moderate (n=2310) | Median [IQR] | 1.0 [0.0, 4.0] | 1.0 [0.0, 2.0] | 1.0 [0.0, 4.0] | 1.0 [0.0, 4.0] | 0.60 | - | 1.0 [0.0, 4.0] | 2.0 [0.0, 4.0] | 1.0 [0.0, 4.0] | 1.0 [0.0, 2.0] | 0.0 [0.0, 0.0] | 2.0 [0.0, 4.0] | <0.001 |
| Functional |  |  |  |  |  |  |  |  |  |  |  |  |  |  |
| mRS (n=2436) | Median [IQR] | 0.0 [0.0, 1.0] | 0.0 [0.0, 1.0] | 0.0 [0.0, 1.0] | 0.0 [0.0, 1.0] | 0.049 | - | 0.0 [0.0, 1.0] | 0.0 [0.0, 0.0] | 0.0 [0.0, 0.0] | 2.0 [0.0, 2.0] | 3.0 [3.0, 3.0] | 4.0 [4.0, 4.0] | <0.001 |
| Post-stroke |  |  |  |  |  |  |  |  |  |  |  |  |  |  |
| Cognition |  |  |  |  |  |  |  |  |  |  |  |  |  |  |
| Cognition, 7-level (n=2436) †‡ | Normal=0 (%) | 1256 (51.6) | 12 (63.2) | 1002 (51.0) | 242 (53.7) | 0.30 | - | - | - | - | - | - | - |  |
|  | Single domain=1 (%) | 530 (21.8) | 3 (15.8) | 441 (22.4) | 86 (19.1) | - | - | - | - | - | - | - | - |  |
|  | Multi-domain=2 (%) | 320 (13.1) | 2 (10.5) | 255 (13.0) | 63 (14.0) | - | - | - | - | - | - | - | - |  |
| Variable |  | All | Severity Unknown | Mild | Severe | p | Severity Unknown | Normal | Single domain | Multi domain | Mild | Moderate | Severe | P for trend |
|  | Mild=3 (%) | 237 (9.7) | 0 (0.0) | 200 (10.2) | 37 (8.2) | - | - | - | - | - | - | - | - |  |
|  | Moderate=4 (%) | 90 (3.7) | 2 (10.5) | 66 (3.4) | 22 (4.9) | - | - | - | - | - | - | - | - |  |
|  | Severe=5 (%) | 3 (0.1) | 0 (0.0) | 2 (0.1) | 1 (0.2) | - | - | - | - | - | - | - | - |  |
|  | Median [IQR] | 0.0 [0.0, 2.0] | 0.0 [0.0, 1.0] | 0.0 [0.0, 2.0] | 0.0 [0.0, 2.0] | 0.57 | - | - | - | - | - | - | - | - |
| Cognition, 4-level (n=2436) ‡ | Normal=0 (%) | 1256 (51.6) | 12 (63.2) | 1002 (51.0) | 242 (53.7) | 0.30 | - | - | - | - | - | - | - |  |
|  | Minor=1 (%) | 850 (34.9) | 5 (26.3) | 696 (35.4) | 149 (33.0) | - | - | - | - | - | - | - | - |  |
|  | Major=2 (%) | 330 (13.5) | 2 (10.5) | 268 (13.6) | 60 (13.3) | - | - | - | - | - | - | - | - |  |
|  | Median [IQR] | 0.0 [0.0, 1.0] | 0.0 [0.0, 1.0] | 0.0 [0.0, 1.0] | 0.0 [0.0, 1.0] | 0.36 | - | - | - | - | - | - | - | - |
| MOCA total (/30) (n=1508) | Mean (SD) | 24.1 (4.3) | 22.0 (5.3) | 24.3 (4.2) | 22.9 (4.5) | <0.001 | - | 25.2 (5.2) | 24.5 (1.8) | 21.4 (2.6) | 24.2 (3.5) | 17.4 (3.6) | - | <0.001 |
| ...<25 (n=1508) | N (%) | 693 (40.6) | 4 (57.1) | 570 (44.3) | 119 (55.9) | 0.0016 | 0 (0.0) | 180 (27.8) | 193 (46.7) | 212 (88.7) | 68 (40.2) | 40 (100.0) | 0 (0.0) | <0.001 |
| T-MOCA (/30) (n=2039) | Mean (SD) | 17.2 (3.7) | 16.4 (3.6) | 17.4 (3.6) | 16.2 (4.0) | <0.001 | - | 17.8 (4.7) | 17.8 (1.2) | 15.2 (2.0) | 17.8 (2.5) | 12.1 (3.1) | 6.0 (3.0) | <0.001 |
| TICS-M total (/37) (n=804) | Mean (SD) | 23.9 (5.1) | 21.4 (4.5) | 24.1 (5.0) | 22.8 (5.3) | 0.0062 | - | 25.5 (6.2) | 24.6 (3.7) | 21.3 (3.4) | 23.3 (3.6) | 17.7 (3.7) | 14.5 (6.4) | <0.001 |
| Verbal fluency (n=2178) | Mean (SD) | 9.9 (4.9) | 9.0 (5.5) | 10.1 (4.9) | 8.6 (4.7) | <0.001 | - | 10.4 (5.3) | 10.5 (4.5) | 8.5 (4.1) | 9.4 (4.5) | 5.9 (4.0) | 3.3 (1.5) | <0.001 |
| Trail making test part A - time (seconds) (n=1645) | Mean (SD) | 67.0 (60.1) | 68.4 (57.5) | 64.2 (56.7) | 82.7 (74.8) | <0.001 | - | 66.0 (63.8) | 59.9 (48.0) | 75.1 (53.6) | 63.2 (37.9) | 111.5 (112.1) | 397.0 (285.7) | <0.001 |
| Trail making test part A - points (n=1645) | Mean (SD) | 23.8 (4.7) | 21.8 (8.8) | 24.0 (4.2) | 22.8 (6.3) | <0.001 | - | 23.8 (4.7) | 24.4 (3.3) | 23.8 (4.7) | 23.8 (5.1) | 20.4 (9.4) | 18.0 (0.0) | <0.001 |
| Trail making test part B - time (seconds) (n=1578) | Mean (SD) | 152.7 (102.6) | 186.5 (126.5) | 150.1 (101.1) | 166.0 (109.2) | 0.029 | - | 141.7 (103.7) | 148.5 (89.3) | 187.9 (110.9) | 154.3 (91.9) | 178.7 (146.9) | - | <0.001 |
| Trail making test part B - points (n=1578) | Mean (SD) | 20.8 (7.4) | 17.6 (11.2) | 21.2 (7.1) | 18.8 (8.7) | <0.001 | - | 20.7 (7.6) | 22.2 (5.7) | 19.8 (8.1) | 21.1 (7.1) | 12.6 (10.1) | - | <0.001 |
| COG-4, from NIHSS [/9] (n=2399) | Median [IQR] | 0.0 [0.0, 0.0] | 2.0 [0.0, 4.0] | 0.0 [0.0, 0.0] | 1.0 [0.0, 2.0] | <0.001 | - | 0.0 [0.0, 0.0] | 0.0 [0.0, 0.0] | 0.0 [0.0, 1.0] | 0.0 [0.0, 0.0] | 1.0 [0.0, 2.0] | 1.0 [1.0, 4.0] | 0.30 |
| **Stroke severity** |  |  |  |  |  |  |  |  |  |  |  |  |  |  |
| NIHSS, worst (/42) (n=2133) | Mean (SD) | 4.9 (4.7) | - | 3.1 (2.0) | 12.3 (4.9) | <0.001 | - | 4.9 (4.6) | 4.6 (4.2) | 5.2 (5.2) | 4.9 (5.0) | 6.3 (5.2) | 13.5 (9.2) | 0.0075 |
| Variable |  | All | Severity Unknown | Mild | Severe | p | Severity Unknown | Normal | Single domain | Multi domain | Mild | Moderate | Severe | P for trend |
| ...>7 | N (%) | 417 (19.5) | 0 (0.0) | 6 (0.4) | 411 (95.8) | <0.001 | 0 (0.0) | 220 (19.4) | 80 (17.5) | 59 (21.9) | 38 (18.7) | 19 (26.8) | 1 (50.0) | <0.001 |
| ...>12 | n (%) | 154 (7.2) | 0 (0.0) | 0 (0.0) | 154 (35.9) | <0.001 | 0 (0.0) | 83 (7.3) | 31 (6.8) | 17 (6.3) | 13 (6.4) | 9 (12.7) | 1 (50.0) | <0.001 |
| ...>20 | n (%) | 35 (1.6) | 0 (0.0) | 0 (0.0) | 35 (8.2) | <0.001 | 0 (0.0) | 16 (1.4) | 3 (0.7) | 9 (3.3) | 5 (2.5) | 2 (2.8) | 0 (0.0) | <0.001 |
| NIHSS (/42) (n=2380) | Mean (SD) | 2.9 (3.4) | - | 1.9 (1.9) | 7.0 (5.1) | <0.001 | - | 3.0 (3.6) | 2.3 (2.6) | 3.1 (3.4) | 2.5 (2.7) | 4.4 (4.4) | 7.7 (5.0) | <0.001 |
| **Mood** |  |  |  |  |  |  |  |  |  |  |  |  |  |  |
| Zung depression scale (n=985) | Mean (SD) | 46.0 (13.3) | 43.5 (13.9) | 45.3 (13.0) | 49.3 (14.6) | <0.001 | - | 46.2 (14.0) | 44.6 (12.6) | 45.6 (12.2) | 47.3 (13.2) | 50.4 (14.8) | 40.0 (-) | 0.16 |
| Patient health questionnaire (n=1986) | Mean (SD) | 5.3 (5.3) | 3.1 (2.4) | 5.1 (5.1) | 6.4 (5.9) | <0.001 | - | 5.3 (5.3) | 5.0 (4.9) | 5.1 (4.9) | 6.0 (5.9) | 8.0 (6.1) | 6.0 (-) | <0.001 |
| PHQ-2 [/2] (n=2262) | Median [IQR] | 0.0 [0.0, 2.0] | 0.0 [0.0, 1.0] | 0.0 [0.0, 2.0] | 1.0 [0.0, 3.0] | <0.001 | - | 0.0 [0.0, 2.0] | 0.0 [0.0, 2.0] | 0.0 [0.0, 2.0] | 1.0 [0.0, 2.0] | 1.0 [0.0, 3.0] | 2.0 [1.0, 2.0] | 0.36 |
| Generalised anxiety disorder (n=2026) | Mean (SD) | 4.0 (4.8) | 2.9 (3.4) | 3.8 (4.7) | 4.7 (5.3) | 0.0031 | - | 4.1 (4.8) | 3.9 (4.7) | 3.4 (4.4) | 4.2 (5.2) | 5.1 (5.3) | 2.0 (-) | 0.14 |
| **Social support** |  |  |  |  |  |  |  |  |  |  |  |  |  |  |
| Social support survey (n=2293) | Mean (SD) | 17.2 (3.9) | 15.9 (5.0) | 17.2 (3.9) | 17.3 (3.8) | 0.69 | - | 17.2 (3.9) | 17.2 (3.7) | 17.2 (3.9) | 17.2 (3.9) | 17.2 (4.1) | 16.7 (5.8) | 1.00 |
| **Frailty** |  |  |  |  |  |  |  |  |  |  |  |  |  |  |
| Clinical frailty score (n=1051) | Median [IQR] | 3.0 [2.0, 4.0] | 3.0 [2.0, 5.0] | 3.0 [2.0, 4.0] | 3.5 [3.0, 5.0] | <0.001 | - | 3.0 [2.0, 4.0] | 3.0 [2.0, 3.0] | 3.0 [2.0, 4.0] | 3.0 [2.0, 4.0] | 5.0 [4.0, 6.0] | 6.0 [6.0, 6.0] | <0.001 |
| **Fatigue** |  |  |  |  |  |  |  |  |  |  |  |  |  |  |
| Brief Fatigue Inventory, unusually tired (n=2070) | Yes (%) | 1182 (57.1) | 7 (77.8) | 944 (55.4) | 231 (64.7) | 0.0012 | 0 (0.0) | 570 (57.4) | 270 (54.5) | 175 (58.7) | 130 (59.1) | 36 (57.1) | 1 (100.0) | <0.001 |
| **Informants** |  |  |  |  |  |  |  |  |  |  |  |  |  |  |
| Number of Informants, expected | N | 674 | 6 | 520 | 148 |  | 0 | 324 | 142 | 82 | 78 | 45 | 3 |  |
| Number of Informants, actual | n (%) | 665 (97.8) | 6 (100.0) | 514 (97.9) | 145 (97.3) |  | 0 (0.0) | 316 (96.3) | 141 (97.9) | 82 (100.0) | 78 (100.0) | 45 (100.0) | 3 (100.0) |  |
| **Informant Outcomes** |  |  |  |  |  |  |  |  |  |  |  |  |  |  |
| IQCODE (n=535) | Mean (SD) | 3.1 (0.5) | 3.6 (1.1) | 3.2 (0.5) | 3.1 (0.5) | 0.39 | - | 3.1 (0.5) | 3.1 (0.4) | 3.2 (0.3) | 3.1 (0.6) | 3.7 (0.7) | 3.0 (0.0) | <0.001 |
| 4AT - Change in alertness (n=613) | Yes (%) | 141 (23.0) | 2 (50.0) | 105 (22.1) | 34 (25.4) | 0.43 | 0 (0.0) | 67 (23.1) | 29 (23.0) | 12 (16.0) | 14 (18.4) | 18 (41.9) | 1 (33.3) | <0.001 |
| Variable |  | All | Severity Unknown | Mild | Severe | p | Severity Unknown | Normal | Single domain | Multi domain | Mild | Moderate | Severe | P for trend |
| Neuropsychiatric Inventory, Questionnaire - Apathy (n=607) | Yes (%) | 152 (25.0) | 3 (75.0) | 117 (24.8) | 32 (24.4) | 0.93 | 0 (0.0) | 72 (25.0) | 23 (18.4) | 14 (18.9) | 15 (19.7) | 27 (65.9) | 1 (33.3) | <0.001 |
| Pre-stroke mRS (n=511) | Median [IQR] | 0.0 [0.0, 2.0] | 1.0 [0.0, 3.0] | 0.0 [0.0, 2.0] | 0.0 [0.0, 2.0] | 0.88 | - | 0.0 [0.0, 1.0] | 0.0 [0.0, 1.0] | 0.0 [0.0, 1.0] | 1.5 [0.0, 2.0] | 3.0 [3.0, 3.0] | 4.0 [3.0, 4.0] | <0.001 |
| Clinical Frailty, /7 (n=503) | Median [IQR] | 3.0 [2.0, 4.0] | 4.0 [2.0, 5.0] | 3.0 [2.0, 4.0] | 3.0 [2.0, 5.0] | 0.27 | - | 3.0 [2.0, 3.0] | 3.0 [2.0, 3.0] | 3.0 [2.0, 3.0] | 3.0 [2.0, 4.0] | 6.0 [5.0, 6.0] | 7.0 [7.0, 7.0] | <0.001 |
| Barthel Index (n=470) | Mean (SD) | 93.4 (15.4) | 100.0 (0.0) | 94.3 (13.6) | 89.8 (20.8) | 0.011 | - | 95.3 (13.1) | 97.3 (6.1) | 93.4 (14.8) | 94.6 (13.9) | 71.4 (24.0) | 27.5 (10.6) | <0.001 |
| Lawton's ADL (n=421) | Mean (SD) | 12.4 (6.4) | 8.3 (0.6) | 12.4 (6.5) | 12.7 (6.3) | 0.67 | - | 11.1 (5.1) | 10.3 (4.3) | 11.0 (5.1) | 12.7 (5.6) | 24.6 (6.3) | 23.5 (6.4) | <0.001 |

† 7-level cognitive outcome comprises: normal, single domain minor neurocognitive disorder, multi-domain minor neurocognitive disorder, minor dementia, moderate dementia, severe dementia.
‡ Death excluded from baseline.
ADL: activities of daily living; EQ-5D-5L HSUV: EuroQoL-5 Dimension-5 level Health Status Utility Value; IQCODE: Informant Questionnaire on Cognitive Decline in the Elderly; MoCA: Montreal Cognitive Assessment; mRS: modified Rankin scale; NIHSS: National Institutes of Health stroke scale; PHQ-2: patient health questionnaire-2; PHQ-9: patient health questionnaire-9; TICS-M: telephone interview cognition scale-modified; t-MOCA: telephone MoCA; 4AT: 4 A's Test (Alertness, Abbreviated mental test-4, Attention and Acute/fluctuating change).

Most participants had an admission CT scan, 2329 (95.6%) and many also had MRI brain imaging, 1118 (45.9%) (Table 3). As reported by the recruiting site, an acute stroke lesion was present in 1727 (71.1%) participants with evidence of mass effect in 186 (7.7%). Of pre-stroke features, there was atrophy in 556 (23.2%), WMH in 665 (27.8%) and evidence of previous stroke in 662 (27.5%). On carotid imaging, stenosis >=50% was present on the left in 121 (7.7%) and right in 128 (8.2%) of participants. Baseline ECG showed AF in 398 (16.6%) (Table 3).

**Table 3.** Baseline investigations by stroke severity (NIHSS≤7 vs >7) and cognitive status. Data are number (%), median [interquartile range] or mean (standard deviation). Comparison by Chi-square, Mann-Whitney U test and t-test or Jonckheere-Terpstra (JT), Kendall’s Tau and ANOVA for trend

| Baseline |  | Patients | Stroke | Severity |  |  | Cognition | Cognition |  |  |  |  |  |  |
| --- | --- | --- | --- | --- | --- | --- | --- | --- | --- | --- | --- | --- | --- | --- |
| Variable |  | All | Unknown | Mild | Severe | p | Unknown | Normal | Single domain | Multi domain | Mild | Moderate | Severe | p |
| Number of patients | N | 2437 | 20 | 1966 | 451 |  | 1 | 1256 | 530 | 320 | 237 | 90 | 3 |  |
| **Investigations** |  |  |  |  |  |  |  |  |  |  |  |  |  |  |
| Type of scan after index event (n=2437) | CT (%) | 2329 (95.6) | 17 (85.0) | 1866 (94.9) | 446 (98.9) | <0.001 | 0 (0.0) | 1194 (95.1) | 500 (94.3) | 312 (97.5) | 230 (97.0) | 90 (100.0) | 3 (100.0) | <0.001 |
|  | MRI (%) | 1118 (45.9) | 4 (20.0) | 993 (50.5) | 121 (26.8) | <0.001 | 0 (0.0) | 615 (49.0) | 248 (46.8) | 127 (39.7) | 101 (42.6) | 27 (30.0) | 0 (0.0) | <0.001 |
| Acute stroke lesion present (n=2428) | Yes (%) | 1727 (71.1) | 16 (84.2) | 1362 (69.6) | 349 (77.4) | <0.001 | 0 (0.0) | 936 (74.9) | 371 (70.1) | 216 (67.5) | 141 (59.5) | 61 (67.8) | 2 (66.7) | <0.001 |
| ...Side of brain (n=1727) | Left (%) | 803 (46.5) | 3 (18.8) | 661 (48.5) | 139 (39.8) | 0.0036 | 0 (0.0) | 432 (46.2) | 178 (48.0) | 99 (45.8) | 62 (44.0) | 31 (50.8) | 1 (50.0) | <0.001 |
|  | Right (%) | 924 (53.5) | 13 (81.3) | 701 (51.5) | 210 (60.2) | 0.0036 | 0 (0.0) | 504 (53.8) | 193 (52.0) | 117 (54.2) | 79 (56.0) | 30 (49.2) | 1 (50.0) | <0.001 |
| Evidence of mass effect (n=2406) | Yes (%) | 186 (7.7) | 2 (11.8) | 118 (6.1) | 66 (14.8) | <0.001 | 0 (0.0) | 95 (7.7) | 39 (7.4) | 28 (8.9) | 15 (6.4) | 8 (9.0) | 1 (50.0) | <0.001 |
| Evidence of atrophy (n=2398) | Yes (%) | 556 (23.2) | 6 (35.3) | 440 (22.6) | 110 (25.2) | 0.24 | 0 (0.0) | 250 (20.3) | 117 (22.4) | 81 (25.6) | 77 (32.8) | 31 (34.8) | 0 (0.0) | <0.001 |
| Evidence of WMH (n=2393) | Yes (%) | 665 (27.8) | 5 (31.3) | 549 (28.3) | 111 (25.4) | 0.22 | 0 (0.0) | 312 (25.3) | 142 (27.2) | 95 (30.2) | 77 (32.8) | 38 (43.2) | 1 (50.0) | <0.001 |
| Evidence of old stroke (n=2406) | Yes (%) | 662 (27.5) | 7 (38.9) | 544 (28.0) | 111 (25.1) | 0.21 | 0 (0.0) | 321 (25.9) | 135 (25.8) | 97 (30.9) | 71 (30.0) | 38 (42.7) | 0 (0.0) | <0.001 |
| Brain frailty score [/3] (n=2383) | Median [IQR] | 1.0 [0.0, 1.0] | 1.0 [0.0, 2.0] | 1.0 [0.0, 1.0] | 0.0 [0.0, 1.0] | 0.36 | - | 0.0 [0.0, 1.0] | 0.0 [0.0, 1.0] | 1.0 [0.0, 1.0] | 1.0 [0.0, 2.0] | 1.0 [0.0, 2.0] | 0.5 [0.0, 1.0] | <0.001 |
| Carotid artery assessment (n=2431) | Yes (%) | 1600 (65.8) | 8 (42.1) | 1330 (67.8) | 262 (58.2) | <0.001 | 0 (0.0) | 839 (66.9) | 354 (66.9) | 213 (66.6) | 157 (66.5) | 36 (40.4) | 1 (50.0) | <0.001 |
| ...Type of scan (n=1594) | CTA (%) | 600 (37.6) | 3 (50.0) | 478 (36.0) | 119 (45.6) | 0.022 | 0 (0.0) | 355 (42.4) | 118 (33.4) | 67 (31.6) | 47 (30.3) | 12 (33.3) | 1 (100.0) | <0.001 |
|  | DSA (%) | 7 (0.4) | 0 (0.0) | 7 (0.5) | 0 (0.0) | 0.022 | 0 (0.0) | 2 (0.2) | 3 (0.8) | 2 (0.9) | 0 (0.0) | 0 (0.0) | 0 (0.0) | <0.001 |
|  | MRA (%) | 40 (2.5) | 0 (0.0) | 34 (2.6) | 6 (2.3) | 0.022 | 0 (0.0) | 20 (2.4) | 8 (2.3) | 8 (3.8) | 4 (2.6) | 0 (0.0) | 0 (0.0) | <0.001 |
|  | Ultrasound (%) | 947 (59.4) | 3 (50.0) | 808 (60.9) | 136 (52.1) | 0.022 | 0 (0.0) | 460 (55.0) | 224 (63.5) | 135 (63.7) | 104 (67.1) | 24 (66.7) | 0 (0.0) | <0.001 |
| Variable |  | All | Unknown | Mild | Severe | p | Unknown | Normal | Single domain | Multi domain | Mild | Moderate | Severe | p |
| ...Degree of stenosis - left (n=1563) | <50% (%) | 1411 (90.3) | 5 (83.3) | 1185 (90.9) | 221 (87.0) | 0.025 | 0 (0.0) | 752 (91.3) | 313 (90.7) | 185 (88.9) | 130 (86.1) | 30 (88.2) | 1 (100.0) | <0.001 |
|  | >=50% (%) | 121 (7.7) | 0 (0.0) | 98 (7.5) | 23 (9.1) | 0.025 | 0 (0.0) | 56 (6.8) | 28 (8.1) | 18 (8.7) | 17 (11.3) | 2 (5.9) | 0 (0.0) | <0.001 |
|  | Occluded (%) | 31 (2.0) | 1 (16.7) | 20 (1.5) | 10 (3.9) | 0.025 | 0 (0.0) | 16 (1.9) | 4 (1.2) | 5 (2.4) | 4 (2.6) | 2 (5.9) | 0 (0.0) | <0.001 |
| ...Degree of stenosis - right (n=1563) | <50% (%) | 1379 (88.2) | 5 (83.3) | 1164 (89.3) | 210 (82.7) | 0.0015 | 0 (0.0) | 726 (88.1) | 308 (89.3) | 185 (88.9) | 132 (87.4) | 27 (79.4) | 1 (100.0) | <0.001 |
|  | >=50% (%) | 128 (8.2) | 1 (16.7) | 101 (7.8) | 26 (10.2) | 0.0015 | 0 (0.0) | 70 (8.5) | 26 (7.5) | 13 (6.3) | 13 (8.6) | 6 (17.6) | 0 (0.0) | <0.001 |
|  | Occluded (%) | 56 (3.6) | 0 (0.0) | 38 (2.9) | 18 (7.1) | 0.0015 | 0 (0.0) | 28 (3.4) | 11 (3.2) | 10 (4.8) | 6 (4.0) | 1 (2.9) | 0 (0.0) | <0.001 |
| Echocardiogram (n=2430) | Yes (%) | 554 (22.8) | 3 (15.8) | 445 (22.7) | 106 (23.6) | 0.67 | 0 (0.0) | 294 (23.4) | 107 (20.2) | 78 (24.5) | 55 (23.3) | 19 (21.3) | 1 (33.3) | <0.001 |
| ...Left ventricular hypertrophy (n=537) | Yes (%) | 99 (18.4) | 0 (0.0) | 81 (18.8) | 18 (17.1) | 0.70 | 0 (0.0) | 56 (19.6) | 15 (14.4) | 13 (16.9) | 13 (25.0) | 2 (10.5) | 0 (0.0) | <0.001 |
| ...Reduced ejection fraction (n=534) | Yes (%) | 42 (7.9) | 1 (50.0) | 31 (7.2) | 10 (9.6) | 0.42 | 0 (0.0) | 18 (6.3) | 9 (8.8) | 4 (5.2) | 8 (15.4) | 3 (15.8) | 0 (0.0) | <0.001 |
| ...Left atrial dilatation (n=537) | Yes (%) | 94 (17.5) | 1 (100.0) | 72 (16.7) | 21 (20.0) | 0.42 | 0 (0.0) | 54 (18.9) | 17 (16.3) | 11 (14.3) | 9 (17.3) | 3 (15.8) | 0 (0.0) | <0.001 |
| ECG (n=2398) | Sinus (%) | 1886 (78.6) | 14 (87.5) | 1551 (79.9) | 321 (72.8) | 0.0037 | 0 (0.0) | 980 (79.4) | 424 (81.7) | 240 (75.9) | 179 (76.2) | 60 (66.7) | 3 (100.0) | <0.001 |
|  | AF (%) | 398 (16.6) | 2 (12.5) | 305 (15.7) | 91 (20.6) | 0.0037 | 0 (0.0) | 192 (15.5) | 68 (13.1) | 63 (19.9) | 50 (21.3) | 25 (27.8) | 0 (0.0) | <0.001 |
|  | Other (%) | 114 (4.8) | 0 (0.0) | 85 (4.4) | 29 (6.6) | 0.0037 | 0 (0.0) | 63 (5.1) | 27 (5.2) | 13 (4.1) | 6 (2.6) | 5 (5.6) | 0 (0.0) | <0.001 |
| ...Left ventricular hypertrophy (n=2309) | Yes (%) | 106 (4.6) | 1 (7.1) | 87 (4.7) | 18 (4.2) | 0.71 | 0 (0.0) | 68 (5.7) | 16 (3.2) | 11 (3.6) | 7 (3.1) | 4 (4.5) | 0 (0.0) | <0.001 |
| Haemoglobin (g/dL) (n=2405) | Mean (SD) | 13.9 (1.9) | 13.6 (1.6) | 13.9 (1.9) | 13.7 (1.9) | 0.0092 | - | 13.9 (1.9) | 14.0 (1.7) | 13.9 (2.2) | 13.7 (2.2) | 13.0 (1.7) | 10.7 (0.9) | <0.001 |
| White cell count ($\times 10^9/L$) (n=2422) | Mean (SD) | 8.6 (5.3) | 13.6 (20.9) | 8.3 (4.3) | 9.3 (7.1) | <0.001 | - | 8.4 (4.4) | 8.9 (8.0) | 8.6 (5.0) | 8.6 (2.8) | 8.0 (2.1) | 11.6 (5.4) | 0.37 |
| Platelets ($\times 10^9/L$) (n=2423) | Mean (SD) | 283.1 (810.0) | 274.9 (53.8) | 272.1 (603.6) | 331.2 (1397) | 0.16 | - | 261.7 (96.3) | 361.8 (1726) | 258.6 (86.1) | 257.3 (102.9) | 269.9 (112.9) | 380.7 (119.7) | 0.27 |
| CRP ($\mu g/dL$) (n=1966) | Mean (SD) | 79.3 (269.1) | 163.8 (274.3) | 78.0 (264.7) | 81.0 (287.1) | 0.85 | - | 77.6 (274.1) | 67.2 (233.0) | 98.7 (289.6) | 92.5 (286.0) | 66.1 (263.8) | 31.7 (48.0) | 0.71 |
| Sodium (mmol/L) (n=2428) | Mean (SD) | 137.6 (13.2) | 137.9 (3.1) | 137.8 (13.0) | 137.0 (14.0) | 0.30 | - | 137.4 (12.6) | 138.2 (16.3) | 137.6 (11.7) | 137.4 (12.5) | 137.9 (3.6) | 134.0 (5.3) | 0.88 |
| Potassium (mmol/L) (n=2381) | Mean (SD) | 4.4 (3.0) | 4.0 (0.4) | 4.4 (2.9) | 4.5 (3.3) | 0.62 | - | 4.3 (2.5) | 4.4 (3.1) | 4.4 (3.3) | 4.2 (0.5) | 5.3 (8.0) | 3.9 (0.5) | 0.079 |
| Variable |  | All | Unknown | Mild | Severe | p | Unknown | Normal | Single domain | Multi domain | Mild | Moderate | Severe | p |
| Glucose (mmol/L) (n=2039) | Mean (SD) | 7.0 (4.4) | 5.3 (2.8) | 7.1 (4.7) | 6.8 (3.3) | 0.30 | - | 7.0 (4.3) | 7.1 (4.8) | 6.7 (4.4) | 7.5 (4.1) | 6.8 (5.1) | 7.3 (2.1) | 0.51 |
| Urea (mmol/L) (n=2314) | Mean (SD) | 6.4 (5.3) | 7.7 (7.9) | 6.4 (5.2) | 6.6 (5.7) | 0.42 | - | 6.5 (5.6) | 6.1 (4.5) | 6.5 (5.3) | 6.7 (4.7) | 7.1 (6.6) | 7.1 (2.3) | 0.58 |
| Creatinine (μmol/L) (n=2426) | Mean (SD) | 83.0 (40.3) | 85.6 (73.8) | 83.8 (37.5) | 79.7 (49.2) | 0.051 | - | 82.5 (45.9) | 83.2 (35.0) | 82.4 (28.9) | 87.6 (36.2) | 80.3 (30.1) | 53.3 (12.2) | 0.38 |
| Estimated or true GFR (mL/min) (n=2412) | Mean (SD) | 73.3 (30.7) | 75.8 (21.3) | 72.8 (29.2) | 75.4 (36.8) | 0.10 | - | 73.5 (27.1) | 76.6 (47.6) | 71.5 (15.1) | 68.8 (16.8) | 70.0 (17.2) | 77.7 (3.1) | 0.020 |

AF: Atrial fibrillation; ECG: electrocardiogram; GFR: glomerular filtration rate;

Overall, 1966 (80.7%) patients were recruited with a TIA or mild stroke (highest NIHSS <=7) and 451 (18.5%) were recruited with a severe stroke (Table 1). Twenty [0.8%] stroke patients did not have NIHSS scores and so severity could not be determined. In comparison with patients with mild stroke/TIA, those with more severe stroke were recruited later, 8.0 [3.0, 17.0] vs 5.0 [2.0-12.0] days, p<0.001. The 7-level ordinal cognition scale did not differ between the groups, median [IQR] 0 [0, 2] vs 0 [0, 2], p=0.57. However, those with severe stroke had worse cognition assessed using individual tests, MoCA 22.9 (4.5) vs 24.3 (4.2), p<0.001; TICS-m 22.8 (5.3) vs 24.1 (5.0), p=0.0062 (Tables 2). Participants with more severe stroke were also more likely to have a visible acute lesion on admission, 348 (77.4%) vs 1362 (69.6%), p<0.001 (Table 3).

Compared with SSNAP data from UK hospitals ^10^ in the same time period (July-September 2019), participants in R4VaD were 5 years younger (median 76 vs 71.0 years), had fewer females (47.6 vs 36.5%), more ischaemic stroke (87.9 vs 92%) and a similar distribution of vascular risk factors and stroke severity (median NIHSS 4.0 vs 4.0).

Blood samples were collected for genome-wide association study (GWAS) analysis from 1159 patients from 31 participating sites. Further blood samples were collected for assessment of inflammatory biomarkers from 407 participants at 14 participating sites. Advanced magnetic resonance imaging (MRI), including diffusion tensor imaging (DTI), was performed in 202 patients at four centres (Aberdeen, Cambridge, Edinburgh, London UCL); this was done as close as possible to the R4VaD early follow up time point.

## DISCUSSION

Post stroke cognitive impairment is common but remains poorly characterised, at least at a level of detail that enables useful advice to be given to individual patients and families. We recruited 2441 participants, more than the planned 2000, and had many more centres than originally planned, this reflecting enthusiasm from NIHR CRN sites to participate. Indeed, many more sites wished to join. We also extended the recruitment period beyond the planned stop date to accommodate recruitment into the MRI substudy which had been affected particularly by Covid-19 restrictions.

The R4VaD study was feasible with 50 sites recruiting participants from 53 who had signed up. These collected detailed baseline data and performed initial follow-up at 6-12 weeks thereby providing evidence that busy stroke centres can deliver cognitive and neuropsychiatric assessments.

Participants received deep phenotyping with multiple demographic, socioeconomic, medical history, clinical, cognition, neuropsychiatric and imaging assessments. As based on MoCA, TICSM and mRS at baseline mapped to DSM-5 diagnostic categories for cognitive impairment using the 7-level ordinal cognition scale, cognition was normal in 51.6%, meaning that 48.4% had single-domain cognitive impairment or worse, amongst whom 13.5% had dementia. Based on the MoCA score alone, 46% had cognitive impairment. Participants with more severe stroke had worse cognition and premorbid mRS at baseline. Since only a very tiny proportion of patients had a history of cognitive impairment (3.6%) or dementia (1.8%) prior to the stroke, most of this substantial burden of cognitive impairment is attributable to the immediate effects of the stroke. Even more concerning, the median stroke severity was neurologically ‘mild’ (NIHSS of 4, identical to the UK patients recorded in SSNAP) underscoring that the focus on neurological impairments has masked the considerable cognitive burden of stroke.

The strengths of the study are its large sample size recruited from a large number of UK sites and deep clinical, cognitive and imaging phenotyping. Weaknesses include a bias towards recruitment of males (59.7%), and limited recruitment during COVID-19 lockdowns. A bias towards recruiting patients with milder stroke follows from our exclusion of patients who were thought unlikely to survive to 6-12 weeks.

Nevertheless, the sample is representative of those surviving the first 12 weeks of stroke in the UK according to SSNAP.

The study is in late follow-up and once this is completed, the database will be cleaned and locked. Therefore, there may be minor changes in the baseline data between those provided here and in the subsequent primary publication. Analysis will follow the statistical analysis plan given here as a supplement. Substudies include advanced neuroimaging, blood biomarker and genetics. The participants have provided consent for being approached regarding future trials. Data sharing plans are given in the SAP.

## FUNDING

R4VaD is funded by the UK Stroke Association, British Heart Foundation and Alzheimer’s Society Priority Programme Award in Vascular Dementia (16 VAD 07) and the UK MRC Dementia Platform UK. The work is also supported by the UK MRC Dementia Research Institute.

### Data availability and sharing

The CI, with approval from the TSC as necessary, will consider all reasonable requests to share individual participant data on provision of a protocol detailing aims, hypotheses, analyses, tables, figures and publication plan. Where possible, we will perform the analyses; alternatively, de-identified data and a data dictionary will be provided for remote analyses, subject to signed data access agreement.

## COMPETING INTERESTS

PB is Stroke Association Professor of Stroke Medicine and an emeritus NIHR Senior Investigator. He has received honoraria from CoMind, DiaMedica, Phagenesis and Roche and has share options with CoMind and DiaMedica.

LJW was funded by the British Heart Foundation (SAVANNAS, PG/19/69/34636). FD is funded by a Stroke Association-Garfield Weston Foundation Senior Clinical Lectureship (TSA LECT 2015/04) and an NHS Research Scotland Research Fellowship.

TQ is funded by a Stroke Association/Chief Scientist Office Scotland Senior Clinical Lectureship for studies in post-stroke cognitive decline. He has received investigator initiated funding from BMS and Pfizer for projects on cardiovascular disease and cognition.

HSM is supported by infrastructure support from the Cambridge University Hospitals NIHR Comprehensive Biomedical Research Centre.

RM has received BP monitoring equipment for research from Omron and is working with them to develop a telemonitoring system. All fees for this work are paid to his institution.

TGR is a NIHR Senior Investigator.

DW has received Honoraria (speaking) from Bayer 2016, 2017, 2018 (talks or debates on ICH, atrial fibrillation, dementia); Honoraria (chairing) from Portola and Bayer (2019); Consultancy fees from Bayer (2017; embolic stroke of undetermined source), Alnylam (2019; CAA), Portola (2019, 2020; andexanet alpha).

APJ is funded by a Stroke Association Margaret Giffen Reader Award (SA L-RC 19\100000).

RT is funded through a British Heart Foundation Chair Award (CH/12/4/29762).

HE is funded by Lancashire Teaching Hospitals NHS Foundation Trust and Lancaster University.

JMW chaired the European Stroke Organisation Guideline Method Working Group on Small vessel Diseases Part 1 Covert SVD and Part 2 Lacunar Ischemic Stroke and the European Stroke Organisation Conference 2021 and 2022

The other authors have no declarations.

