## Supplemental Text, Tables and Figures for "Rates, risks and routes to reduce vascular dementia (R4VaD), a UK-wide multicentre prospective observational cohort study of cognition after stroke: baseline data and statistical analysis plan (ISRCTN18274006)"

### APPENDIX 1. STATISTICAL ANALYSIS PLAN

#### 1. ADMINISTRATIVE INFORMATION

##### 1.1 Title, registration, versions and revisions

***1.1.1 Full study title***: Rates, risks and routes to reduce vascular dementia (R4VaD), a UK-wide multicentre prospective observational cohort study of cognition after stroke: baseline data and statistical analysis plan (ISRCTN18274006)

***1.1.2 Acronym***: Rates, risks and routes to reduce vascular dementia (R4VaD)

***1.1.3 IRAS project number***: 239109

***1.1.4 Registration***: ISRCTN18274006

***1.1.5 Protocol version***: 1.0 (12 Feb 2018)

***1.1.6 SAP version***: 1.0 (30 April 2024)

***1.1.7 SAP revisions***: None

##### 1.2 Roles and responsibilities

***1.2.1 Author***: Philip M Bath

***1.2.2 Responsible statistician***: Lisa J Woodhouse

***1.2.3 Chief Investigator***: Joanna M Wardlaw

***1.2.4 Contributors and roles***:
Philip M Bath: wrote first draft
Rosalind Brown: study manager
Ellen Backhouse: study manager
Lisa J Woodhouse: statistician
Fergus Doubal: refinement of text
Terence J Quinn: refinement of text
Thompson Robinson: refinement of text
Hugh S Markus: refinement of text
Richard McManus: refinement of text
John T O'Brien: refinement of text
David J Werring: refinement of text
Nikola Sprigg: refinement of text
Adrian Parry-Jones: refinement of text
Rhian M Touyz: refinement of text
Steven Williams: refinement of text
Yee-Haur Ma: refinement of text
Hedley Emsley: refinement of text
Joanna M Wardlaw: refinement of text

#### 2. INTRODUCTION

##### 2.1 Background and rationale

The R4VaD study is a large, UK-wide multicentre, longitudinal, representative study of patients presenting with stroke or TIA to Stroke Centres, using standardised proportionate ascertainment methods to assess cognition, functional and neuropsychiatric outcomes up to at least two years after stroke. The protocol has been published.[1]

This Supporting Information Appendix S1 presents the statistical analysis plan (SAP) and a detailed listing of baseline characteristics prior to reporting the primary results. It details the full SAP and is presented prior to locking of the study database so that analyses are not data driven or reported selectively.[2] We also list planned secondary analyses and substudies. The SAP follows the layout suggested by Hiemstra *et al*.[3]

##### 2.2 Objectives

###### *2.2.1 Primary Objective*
To determine rates of cognitive impairment and neurocognitive disorder (including dementia) at up to two years after stroke.

###### *2.2.2 Secondary Objectives*
a) Compare cognition between participants with mild stroke/TIA (highest NIHSS <=7 [4 5]) versus severe stroke (highest NIHSS >7) at baseline; this result is reported here;
b) Compare cognition between participants with mild stroke/TIA (highest NIHSS <=7 [4 5]) versus severe stroke (highest NIHSS >7) at 1 year;
c) Better understand trajectories of cognitive impairment over two years after stroke;
d) Develop and validate a cognition scale for use in stroke patients that is free to use;
e) Identify key risk predictors and develop better risk prediction models for individual patients;
f) Perform studies to improve our ability to undertake cognitive testing after stroke and enhance mechanistic understanding of PSCI;
g) Establish a well phenotyped population, in follow-up, with consent for re-contact for future trials;
h) Provide data to plan future RCTs and services for patients with PSCI.

This SAP focuses on the primary objective. Planned follow-on publications addressing the secondary objectives are listed in section 7.6.

#### 3. STUDY METHODS

##### 3.1 Study design
Prospective observational, longitudinal inception cohort with central follow-up and nested substudies in major UK stroke centres representing geographic and socioeconomic diversity. An electronic case record form (eCRF) [6-9] streamlines baseline and follow-up data collection and verification. Central follow-up by validated telephone and postal methods [6-9] will reduce local research burden, data loss and facilitate analyses. Data collected for UK stroke audits (Sentinel; Stroke National Audit Project [SSNAP [10]], Scottish Stroke care Audit [SSCA]) will determine the study's representativeness of UK hospital-assessed stroke patients. Where appropriate, we

will use trusted research environments/safe havens to link our data to other health datasets to validate outcomes and provide an anonymised, ethics- and governance-approved secure database.

The R4VaD study is intended to offer an enriched resource for future hypothesis generation and hypothesis testing in the field of vascular cognitive impairment. The study has been designed to offer a rich dataset and population available for future research. It would therefore be impossible to detail all the potential analyses that these data could inform. In this paper, we describe the core analyses that are fundamental to address our main research questions regarding the understanding of the epidemiology of post stroke cognitive impairment.

##### 3.2 Sample size/power considerations

Conservatively we have used sample size calculations based on binary measures. Use of ordinal measures will give greater statistical power.

###### 3.2.1 Distribution of cognition

Assuming estimated population prevalence of dementia 0.25, precision 0.025, confidence 0.95 and population size 100,000 (the annual number of new strokes in the UK), we require 1,140 participants (https://epitools.ausvet.com.au/oneproportion). Thus, 2000 patients will give greater confidence.

###### *3.2.2 Comparison of two groups - mild stroke/TIA vs severe stroke (primary analysis)*

Assuming power 0.90 and alpha=0.05, we can detect the following differences in dementia proportion in mild (highest NIHSS <=7 [4,5]) vs. severe (highest NIHSS >7) stroke, respectively: 20% vs. 27% in n=1596, 10% vs. 15% in n=1914, and 10% vs. 20% in n=572 (***S. Table A***). Thus, 2000 patients, with a wide range of stroke severities, will allow us to detect small (absolute difference 5%) but clinically meaningful differences in dementia between mild stroke/TIA vs. severe stroke. This sample size will also be able to detect differences in degrees of VCI and dementia and in subgroups defined by age, pre-morbid cognitive ability, stroke subtype, stroke severity (mild vs. severe) and vascular risk factors. Analysis of ordinal (4- or 7-level ordered categorical scale) will further increase statistical power over binary outcomes.[11,12]

**S. Table A**. Sample size calculation related to rate of dementia by stroke severity: mild versus severe (%)

| Mild stroke/TIA (%) | Severe stroke (%) | Absolute Risk Difference (%) | Sample size, total |
|---|---|---|---|
| 10 | 20 | 10 | 572 |
| 10 | 15 | 5 | 1914 |
| 20 | 40 | 20 | 236 |
| 20 | 30 | 10 | 825 |
| 20 | 27 | 7 | 1596 |
| 20 | 25 | 5 | 3008 |

##### 3.3 Framework

The primary objective is to determine the prevalence and severity of cognitive impairment and neuro-cognitive disorders (including dementia) up to two years after stroke. The primary analyses will compare the distribution of cognition between participants presenting with mild stroke/TIA vs. severe stroke.

##### 3.4 Interim/repetitive analyses and stopping guidance
Data will be tabulated annually for Study Steering Committee meetings. No comparative analyses will be performed until data collection has been completed and the database locked. There are no formal stopping rules, but a failure of recruitment would lead to early study termination.

##### 3.5 Timing of final analyses
These will be performed once data collection has been completed and the database has been locked.

##### 3.6 Timing of outcome assessments
Assessments will be performed at baseline, 6 weeks, 12 and 24 months (***S. Table B***).

**S. Table B**. Assessments at baseline and follow-up by time point

| Activity / assessment | Baseline | 6±2 weeks | 12/24 months | End of study |
|---|---|---|---|---|
| Demographics | X | | | |
| Medical history | X | | | |
| Vital signs | X | | | |
| Premorbid function | X | | | |
| Routine tests [1] | X | | | |
| Vascular events | | X | X | |
| Function | X | X | X | |
| Cognitive testing | X | X | X | |
| Blood sampling | | X | | |
| Genetic sampling [2] | X | | | |
| Neuroimaging [3] | X [4] | | | |
| Data linkage [5] | | | | X |

1. Blood tests, carotid or cardiac imaging, routine diagnostic brain imaging results
2. Genetic substudy - sampling may also be performed at study visit 1
3. Routine diagnostic CT or MRI scanning collected at admission; imaging substudy in some participants - MRI (structural and diffusion tensor)
4. Can be at baseline or 6 weeks
5. Data linkage via NHS Digital and Public Health Scotland to validate our findings and determine long term outcomes up to the end of the study. This includes death, cause of death, further stroke or cardiovascular related events and prescribing information

#### 4. Statistical principles

##### 4.1 Confidence intervals and P-values
The results of analyses and comparisons will be shown with 95% confidence intervals and threshold for determining statistical significance set at $p<0.05$. No adjustment will be made for multiplicity.

##### 4.2 Adherence and protocol deviations
As an observational study with minimal exclusion criteria, we have no listed deviations. All available data will be used without imputation.

##### 4.3 Analysis populations
The sample population will comprise all consented patients in whom at least some baseline data were recorded. Analyses assessing change will include all patients with both baseline and at least some follow-up data collected. Multiple variable analyses will include all patients with complete data for the dependent and each independent variable.

#### 5. Study population

##### 5.1 Screening data
Screening logs will not be kept routinely so that data collection can be prioritised and since the eligibility criteria are wide. A screening log was performed at seven sites for four weeks in Jan – Feb 2020 to assess representativeness of the sample. We will compare the characteristics of the R4VaD sample with SSNAP in: a) all recorded patients in England and Wales, and b) all patients recorded at the R4VaD sites in SSNAP. Data linkage with central health data will be used to validate long term outcomes including COVID infection status.

##### 5.2 Eligibility
To avoid the selection biases of previous studies, and recognising changes in recovery with new treatments, we propose inclusive recruitment:
- Patients aged ≥18 years with no upper limit
- Ischaemic, or spontaneous haemorrhagic (non-traumatic, non-subarachnoid haemorrhage, non-AVM) stroke or transient ischaemic attack (TIA) within 6 weeks of onset
- Expected to survive at least to 12 weeks post stroke

Due to the varying and evolving concepts and definitions of PSCI and VaD, and to avoid assumption bias, we will not restrict the cohort but rather will include as many patients of all severities and subtypes as possible, regardless of their potential post stroke cognitive syndromes. We will collect the necessary investigations, cognitive and functional data to allow varying diagnostic criteria and stratification to be explored. Participants who are not deemed to have entered the study with cerebrovascular event (ICH, IS, TIA) will be excluded from all analyses.

##### 5.3 Recruitment
Recruitment will be summarised in a STROBE flow diagram.[13 14]

##### 5.4 Withdrawal/follow-up
Withdrawals and missed follow-ups and their timing will be summarised in the STROBE flow diagram.

##### 5.5 Baseline patient characteristics
Baseline characteristics will comprise demographic, education, premorbid function, medical history, clinical parameters and in-hospital reperfusion treatments (a revised version of Table 1 in the linked Baseline paper). Data on COVID-19 exposure and treatment have been collected since April 2020.

##### 5.6 Confounding covariates
The majority of measured variables are likely to be correlated as most relate to the cognitive status of a patient. Example confounding variables are given and these are categorised by whether these were 'measured', as per routine practice, or 'unmeasured'; the latter will lead to residual confounding. We have listed those variables known to have the strongest association with PSCI but given the relative

lack of knowledge in the area it is plausible that our study may suggest new, important risk factors.

##### *5.6.1 Cognition*

*5.6.1.1 Measured*

Age, highest education, main occupation, socioeconomic status, stroke type and severity, small vessel disease, mood, premorbid function, pre-stroke dementia, diabetes mellitus, post-stroke complications, social isolation, vision and hearing.

*5.6.1.2 Unmeasured*

An example is cardiac function.

##### *5.6.2 Mortality*

5.6.2.1 Measured

Age, comorbidities, stroke severity.

5.6.2.2 Unmeasured

Cause of mortality, patient/family wishes regarding extent of treatment/prevention.

#### 6. Analysis

##### 6.1 Outcome

##### *6.1.1 Primary outcome*

The primary outcome is a seven-level ordered categorical scale (Figure 1) measured at 2 years:[1]

1. normal cognition
2. mild neurocognitive disorder with impairment in one cognitive domain
3. mild neurocognitive disorder with impairment in two or more domains
4. major neurocognitive disorder or dementia, mild
5. major neurocognitive disorder or dementia, moderate
6. major neurocognitive disorder or dementia, severe
7. death (the inevitable consequence of severe dementia)

The scale operationalises the DSM-5 categories with additional clinically- and participant-relevant information driven by information from our core data set:

- Montreal Cognitive Assessment (MoCA) or Telephone Interview for Cognitive Status (TICS) [15 16]
- modified Rankin Scale (and/or Barthel index) [17 18]
- IQCODE, disposition (need for care-home)
- evidence of clinical dementia diagnosis according to a validated adjudication process (which will include but not be restricted to formal diagnosis, taking a cholinesterase inhibitor or memantine) or death

The operationalisation of neurocognitive disorder is dependent on data from cognitive assessments in combination with data on function, where the distinction between minor and major disorder is based on presence of cognitive impairment and evidence of functional deficits. The threshold scores used to signify impairments in cognition and function are based on usual reference points for the scales when applied in stroke.[19] Where there is no agreed threshold in stroke, for example the cognitive score to signal transition from moderate to severe cognitive impairment, then thresholds were taken from the broader dementia test literature (https://www.mocatest.org/faq/). In general, for the operationalisation of moderate and severe, we used ability in extended and basic activities of daily living as well as absolute score on cognitive testing. The operationalisation was discussed and amended by the study applicants and further revised by the steering group until consensus was reached.

The seven-point scale is an expanded, operationalised version of a four-point scale which reflects the categories of the DSM-5 neurocognitive disorder classification (normal, mild, major), while encompassing practical factors that are common in stroke such as functional impairments. Therefore, the seven-point scale has practical granularity of patient-relevant diagnoses but can also be condensed and expressed in terms of internationally agreed dementia diagnostic categories. However, we believe that the seven levels offer a more nuanced description of important cognitive states that are not distinguished in the four level or DSM 5 categories.[1] The distinction of various levels of severity within the major neurocognitive disorder rubric is especially pertinent as in clinical practice and trials, dementia is often categorised as mild, moderate and severe.

We recognise that our operationalisation, while based on valid principles, is a novel approach. We have embedded validation steps within the study program. Validation will be based on expert adjudication of clinical and cognitive data. These processes will be fully described in a separate manuscript. Our secondary analyses will use more traditional outcome measures such as change in cognitive score and incident clinical diagnosis. We will assess for consistency across our novel and traditional metrics.

##### 6.1.2 Secondary outcomes - participant

Each outcome domain comprises the key measure (given first) and then others in order of priority of data collection. Measures will be made at baseline and then at 6 weeks, 12 and 24 months.

*6.1.2.1 Death*

In patients who have died, a worst value will be assigned for each scale,[8 20-22] an approach that means patients who have died are not lost from analyses, anchors the scores to each other, reduces bias, and increases statistical power. Death is considered worse than any living score since it cannot be reversed whereas any living status could, potentially, be improved by future treatments. The following lists the scales, the abbreviation and range from best to worst score, followed by the score value for death.

Death will be analysed as:

- Time to all cause
- Cause-specific: dementia, stroke, myocardial infarction, infection, other

*6.1.2.2 Raw data versus z scores*

It is traditional in neuropsychological testing to take raw scores on tests and standardise using age, sex, education matched normative values and describe as z scores. Definitions of mild and major neurocognitive disorder have been proposed based on scores between 1-2 standard deviations and below 2 standard deviations, respectively. Our primary analyses will use raw scores, rather than normed values. This recognises that contemporary, culturally relevant norms are not available for all the tests included in our battery and that analysis is possible using unadjusted data. We will correct for age, sex, education and other cognitive confounders in our planned comparative analyses.

*6.1.2.3 Cognition (depending on whether in person or remotely collected)*

- 4-level cognition/dementia scale (3 to 1, death = 0) (Figure 1)
- Montreal cognition assessment (MoCA, 30 to 0, death = -1) [23 24]
- Trails A score (25 to 0, death = -1) [25]
- Trails A time (0 to maximum time, death = max. time +1)
- Trails B score for first 10 points (25 to 0, death = -1)
- Trails B time (0 to maximum time, death = max. time +1)
- Telephone interview of cognition scale-modified (TICS, 39 to 0, death = -1) [15 26]
- Letter digit coding (LDC, no maximum to 0, death = -1) [27]
- Hopkins verbal learning test (HVLT) [28]
- Boston naming test (BNT, no maximum to 0, death = -1) [29]
- Verbal fluency phonemic (letters A/F/S, maximum to 0, death = -1) [30]
- Memory or thinking problem (0 to 1, death = 2)
- Dementia, clinical diagnosis, e.g. from memory clinic (0 to 1, death = 2)
- Dementia, adjudicated diagnosis (substudy) (0 to 1, death = 2)

*6.1.2.4 Activities of daily living (ADL)*
- Barthel index (BI, 100-0, death = -5) [31 32]
- Lawton ADL (LADL, 8 to 0, death = -1) [33]

*6.1.2.5 Fatigue*
- Brief fatigue inventory (binary BFI, 0 to 1, death = 2) [34]
- Brief fatigue inventory (continuous BFI, 0 to 90, death = 91) [34]

*6.1.2.6 Frailty*
- Clinical frailty score (CFS, 1 to 9, death = 10) [35]

*6.1.2.7 Functional Outcome*
- modified Rankin Scale (mRS, 0 to 5, death = 6) [17 18]
- Disposition (home/residential care/nursing care/hospital/death)

*6.1.2.8 Mood*
- Patient health questionnaire-2 (PHQ-2, 0 to 1, death = 2) [36]
- Patient health questionnaire-9 (PHQ-9, 0 to 27, death = 28)
- Generalised anxiety disorder (GAD-7, 0 to 21, death = 22) [37]
- Zung depression scale (ZDS, 0 to 100, death = 102.5) [38]
- Office National Statistics-personal well-being 4-item (ONS-4, 40 to 0, death = -1) [39]
- Depression, clinical diagnosis (0 to 1, death = 2)

*6.1.2.9 Quality of life*
- Euro-quality of life-5 dimension-5 level (EQ-5D-5L, 1 to -0.5, death = 0) [40]
- Euro-quality of life-visual analogue scale (EQ-VAS, 100 to 0, death = -1)

*6.1.2.10 Social Support*
- Medical Outcomes Study Social support survey (MOS-SSS-4 20 to 4, death = 3) [41]

*6.1.2.11 Socioeconomic status (SES)*
- Occupation, postcode (current SES)
- Father’s and mother’s occupation (childhood SES)

*6.1.2.12 Global*
- Stroke impact scale, includes domains of: ADL, communication, emotion, hand function, handicap, memory, mobility, strength (SIS, 59 to 0, death = -1) [42]

##### 6.1.3 Secondary outcomes - Informant

*6.1.3.1 Informant scoring*

- Informant Questionnaire on cognitive decline in the elderly (IQCODE) [43]
- Delirium (4AT, 0 to 12, death = 13) [44]
- Neuropsychiatric Inventory (NPI-Q) [45]
  - Total items (0 to 12, death = 13)
  - Total severity (0 to 36, death = 37)
  - Total distress (0 to 60, death = 61)
- Zarit burden interview (0 to 88, death = 89) [46]
- Clinical frailty score (CFS, 1 to 9, death = 10) [35]
- modified Rankin Scale (mRS, 0 to 5, death = 6) [18]

#### 6.2 Analysis methods

##### 6.2.1 Analysis of outcomes

*6.2.1.1 Primary outcome*

The primary outcome will be presented numerically, as median [interquartile range], and graphically. When comparing the primary endpoint between participants with mild stroke/TIA vs. severe stroke, the distribution across all 7 levels will be assessed using ordinal logistic regression (OLR, using SAS PROC GENMOD using ordinal approaches improves statistical power.[11 12]

*6.2.1.2 Analyses of secondary outcomes*

Key secondary analyses will comprise:

- Comparison of the distribution of cognitive impairment at year 2 versus baseline using ordinal logistic regression.
- Trajectory of change in cognitive impairment over 2 years using ordinal repeated measures (using SAS PROC GENMOD with REPEATED statement).

Central tendency, comparisons and regressions will be analysed as follows (***S. Table C***).

**S. Table C. Descriptive and analytical statistics**

| | Binary | Nominal | Ordinal | Continuous |
|---|---|---|---|---|
| Central tendency and distribution | N (%) | N (%) | Median [interquartile range] | Mean (standard deviation) |
| Comparisons | Chi-square (2x2) | Chi-square (2x2, or rxc) | Mann-Whitney U (MWU) or Kruskal-Wallis (KW) | t-test (pooled) or 1-way ANOVA |
| Regression | Binary logistic regression | - | Ordinal logistic regression (OLR) | Multiple linear regression (MLR) |
| Trend | Binary repeated measures (BRM) | - | Ordinal repeated measures (ORM) | Continuous repeated measures (CRM) |
| Umbrella alternatives | - | - | Mack-Wolfe umbrella, peak unknown (MWUA) [47 48] | MWUA |

##### 6.2.2 Covariate adjustment

Analyses will be adjusted for baseline covariates where possible:

- Clinical: age (raw data), sex (female, male), educational attainment (primary/secondary [age~<16 years] vs GCSE/O'level/Scottish lower [age~16] vs A'level/Scottish higher [age~18] vs under/post-graduate university), pre-stroke mRS (individual levels), stroke type (TIA, IS, ICH), stroke severity (NIHSS, raw data), 7-level cognition and time from stroke onset to consent/assessment.
- Neuroimaging: acute stroke findings (lesion size, location); atrophy (global or and hippocampal), white matter hyperintensities (WMH), prior infarcts, SVD score, frailty score.

Covariate adjustment with ordered categorical (mRS, educational attainment) or continuous (age, NIHSS, time from stroke onset to consent) variables will use original, not dichotomised, data.

##### 6.2.3 Assumption checking

The assumption of proportionality will be tested using the likelihood ratio test.

##### 6.2.4 Alternative methods

If the data fail the assumption of proportionality (tested using the likelihood ratio test), we will use multiple linear regression instead since the analysis will be based on 7 levels and a large number of patients (central limit theorem).

##### 6.2.5 Sensitivity analyses

In addition to assessment of raw data, the primary outcome will be compared between mild stroke/TIA vs. severe stroke using additional statistical approaches in sensitivity analyses:

- Unadjusted comparison: ordinal logistic regression
- Dichotomous comparison: binary logistic regression of normal versus cognitive impairment/dementia/death (adjusted)
- Continuous repeated measures (adjusted)
- With imputation of missing data using multiple regression imputation: ordinal logistic regression (adjusted) [49]

##### 6.2.6 Analysis of primary outcome in subgroups

The 7-level will be studied in pre-specified subgroups comprising baseline variables, both those used in adjustment and other likely prognostic factors:

- Clinical adjustment variables: age (<70 v >=70 years), sex (female v male), educational attainment (as per section 6.2.2), pre-stroke mRS (0 v >0), stroke type (TIA v IS v ICH), stroke severity (NIHSS 0-3, 4-7, >7) and time from stroke onset to consent/assessment
- Neuroimaging adjustment variables: atrophy (global, hippocampal), white matter hyperintensities (WMH), prior infarcts, SVD score, frailty score
- Other clinical variables: history of hypertension, history of stroke, systolic blood pressure at initial assessment, stroke syndrome (OCSP [50]), side of signs (right, bilateral, left)
- Other neuroimaging variables: acute lesion visible, lesion size, location
- Socioeconomics: main occupation, deprivation index (from postcode, https://census.ukdataservice.ac.uk/get-data/related/deprivation.aspx)

The results of these subgroup analyses will not be adjusted for multiple testing. These analyses are intended to be hypothesis generating as the study is not powered to detect differences in individual subgroups. Other factors may be used to adjust for analyses of secondary outcomes.

##### 6.3 Missing data

Missing data may occur at outcome level or at test level or within a test at component item level. Notably, some tests have to exclude components if performed by telephone and/or postal questionnaire. There is often a relationship between inability to complete outcome assessment and cognitive function or neurological deficit after stroke (e.g. inability to hold a pen) and so assumptions around random missingness may not be valid, even if the patterns of missing data initially suggest 'missing completely at random' status. Indeed, failure to complete a test may be an indicator of cognitive impairment rather than real missingness.

The approaches taken to missing cognitive data can have a substantial effect on epidemiological estimates.[49] We will use the approach that makes greatest use of available data.

We have permissions to allow for linkage of the study dataset to primary and secondary care electronic health records. This will allow for an assessment of incident cognitive syndrome and other clinical outcomes and medications across all the participants. Evidence of dementia diagnosis coming from data linkage will supersede data collected during the study.

##### 6.4 Additional analyses - Global analyses

We will integrate multiple scores into one global analysis and so provide a more holistic comparison which also improves statistical power:

- Global, overall: 7-level cognition scale, mRS, BI, PHQ-9, EQ-5D-5L HUS
- Global, cognition: 7-level cognition scale, verbal fluency
- Global, mood: PHQ-9, GAD, ZDS

Analyses will use the Wei-Lachin test [51-54] with comparison of data at 2 years with baseline.

Other global scores will be calculated:

- Global physical/mobility as sum of mobility measurements in different scales = EQ-5D mobility + BI get about
- Global neuro-psychiatric status as sum of measures of anxiety, depression, fatigue and apathy = GAD + PHQ-9 + BFI + NPI (informant)

##### 6.5 Statistical software

SAS version 9.4 (SAS Institute, Cary, NC, USA).

#### <u>7. Additional information</u>

##### 7.1 Governance

R4VaD is approved by Ethics Committees in England (Health Research Authority), Wales (Health and Care Research, Newcastle), Northern Ireland (all Northeast Newcastle and North Tyneside 1; Ref number 18/NE/0150), Scotland (A Research Ethics Committee; Ref number 18/SS/055). NHS Research and Development/Innovation approval is given in each participating site. The study is adopted by the National Institute for Health and social care Research (NIHR) Clinical Research Network in England, Wales and Northern Ireland, and the Stroke Research Network in Scotland.

##### 7.2 Minimising bias

Multiple approaches will be taken to minimise bias: central data registration with real-time on-line validation; blinded central postal and/or telephone assessment of outcomes; blinded adjudication of neuroimaging; inclusion of patients enrolled in other studies (co-enrolment); analysis by pre-specified subgroups; and analysis by intention-to-enrol (i.e. all participants).

##### 7.3 Discrepancies between protocol and statistical analysis plan

Where there is a difference between the protocol (on website), published protocol [1] and SAP, the SAP will take precedence.

##### 7.4 Calculations

###### 7.4.1 Brain frailty and SVD scores

Based on neuroimaging:

- Brain frailty = Atrophy + WML + Previous stroke lesion
- SVD score = WML, lacunes (a full score can be calculated where MRI is available)

###### 7.4.2 Cognitive domains, based on DSM-5 [55]

We will calculate scores for cognitive domains using sub-scores of MoCA (or TICS if missing). We recognise that in global cognitive assessments it can be difficult to map a test item to a single cognitive domain, for example the clock drawing test in the MoCA includes aspects of attention, executive function and visual-perceptual function.

- Learning and memory: orientation in place (from MoCA), delayed recall of five word (MoCA), and recall and delayed recall of ten words (TICS)
- Language: using comprehension, semantic and recent memory (from MoCA; similar elements in TICS-M)
- Perceptual-motor function: Cube copy and clock drawing from MoCA
- Executive function: Trail making tests A & B, verbal fluency test (VFT-phonemic)-F (from MoCA); verbal fluency test (VFT-semantic)-animals; clock drawing test (from MoCA); digits forward (from MoCA); digits backward (from MoCA)
- Complex attention: using serial sevens subtraction (MoCA), letter tapping (MoCA)
- Social cognition is not classically assessed in cognitive screening tools and there are no agreed generic short form assessments for social cognition. Aspects of social cognition will be assessed through informant data and NPI-Q although these are not part of the core outcome set.

##### 7.5 Montreal cognitive assessment-modified (MoCA-m) trails

MoCA trail is not always collected but rather estimated from the Trails B score (where available):

- If Trails B score <12 then MoCA trail = 0
- If Trails B score >=12 then MoCA trail = 1

Prior to participant 475, MoCA trail will be provided for participants recruited in Edinburgh and for others it will be calculated from the Trails B score. From participant 475 onwards, it will be determined from the Trails B accuracy on the first 10 points.

##### 7.6 Stroke subtype according to Oxfordshire Community Stroke Project (OCSP)

The OCSP will not be collected but will be derived from the NIHSS:[56]

1. Hemianopia from visual fields =2
2. Weakness = (motor arm>0) OR (motor leg>0) OR (facial palsy>0)
3. Sensory >0
4. Cortical = (language>0) OR (visual inattention=1) OR (sensory inattention=1)

LACI = (2 OR 3) NOT (1 OR 4)
TACS = 1 AND (2 OR 3) AND 4
POCS = ATAXIA OR 1 NOT 4
PACS = NOT LACI OR TACS OR POCS

##### 7.7 Publications, published and planned

1. Protocol - published [1]
2. SAP and baseline data - this publication
3. Early outcomes – Stroke severity, dependency, cognitive and mood outcomes at 6 weeks post-stroke.
4. Dementia adjudication vs dementia diagnosis – Comparison of dementia adjudication process and assessments in the main database in a subsample of participants (n=250)
5. Primary results paper – rates, levels, risk factors for and trajectories of cognitive impairment at up to 2 years after stroke
6. Practicalities of tests *and implications for cognitive status* based on completeness vs NIHSS, vision, deafness, mRS etc; simpler v longer cognitive tests – MoCA, TICS v letter digit coding, Boston Naming, CERAD, verbal fluency (A and S)
7. Development of a free-to-use cognition scale
8. Relationship between cognition and BP
9. Validation of simple imaging markers including acute lesion location and their contribution to prediction of cognitive impairment including comparison of site and central adjudication
10. Analysis of outcomes to improve efficiencies in future research
    i. Inferential statistics versus machine learning algorithms
    ii. Components of scales to create novel mobility scales
11. Biomarker/inflammation substudy
12. Genetic markers substudy
13. Diffusion tensor imaging and MRI findings substudy
14. Other publications as determined by the Study Steering Committee

##### 7.8 Data sharing

In the future, the anonymised study data will be made available for use by external investigators in appropriate analyses upon request via a publicly accessible portal (e.g. University of Edinburgh data share). Data from R4VaD will also be shared as appropriate with individual patient data pooling projects involving stroke and dementia; a non-inclusive list includes:

- Dementia Platform UK data portal (https://www.dementiasplatform.uk/)
- Virtual International Stroke Trials Archive-Cognition (VISTA-COG)
- Virtual International Cardiovascular and Cognitive Trials Archive (VICCTA, http://www.virtualtrialsarchives.org)
- STROKOG (https://cheba.unsw.edu.au/consortia/strokog)
- META-VCI Map (https://metavcimap.org/)

Similarly, anonymised neuroimaging data will be published.[57] The mechanisms and processes for managing external access will be determined during the course of the study. Proposals will be considered by the R4VAD Sub studies committee.

##### 7.9 Substudies

This SAP does not cover a number of substudies which will have their own analysis plans:

- Diffusion tensor imaging: Examination of a range of DTI metrics and other neuroimaging variables on cognitive, functional and neuropsychiatric outcomes.
- Dementia adjudication: Comparison of local site dementia assessment versus that collected for the main database.
- Genotyping: Cross-sectional and cohort phenotypic changes by genotype

**Table 1. Main Outcomes in all participants.** (The supplement will contain the same table in only participants with complete data.)

Data are number (%), median [interquartile range] or mean (standard deviation). Comparisons of outcomes at times 0-2, 6, 52 and 104 weeks by Mack-Wolfe umbrella alternatives test, peak unknown (MWUA). Comparisons at two time points by Chi-square test (CST) or Mann-Whitney U test (MWU). Comparisons of outcomes in mild v severe stroke at 2 year by binary logistic regression (BLT), binary repeated measures (BRM), ordinal logistic regression (OLR) or multiple linear regression (MLR) with adjustment for baseline value, age, sex, mRS at baseline, stroke type (ICH, IS, TIA) and time from stroke onset to baseline. Global analyses used the Wei-Lachin test (WLT [51-54]).

| **Week** | **0 (0-6)** | **6 (4-8)** | **52** | **104** | **Peak (wk)** | **p** | **Mild stroke/TIA** NIHSS <=7 | **Severe stroke** NIHSS>7 | **Differ-ence (95% CI)** at 2 years | **p** |
|---|---|---|---|---|---|---|---|---|---|---|
| Number | xxxx | xxxx | xxxx | xxxx | | | xxxx | xxxx | | |
| **Cognition, participant Primary, all data** | | | | | | | | | | |
| Cognition - 7 level | xx.x [xx.x,xx.x] | xx.x [xx.x,xx.x] | xx.x [xx.x,xx.x] | xx.x [xx.x,xx.x] | | MWUA | xx.x [xx.x,xx.x] | xx.x [xx.x,xx.x] | | aOLR |
| Normal | xxxx (xx.x) | xxxx (xx.x) | xxxx (xx.x) | xxxx (xx.x) | | | xxxx (xx.x) | xxxx (xx.x) | | |
| Single-domain | xxxx (xx.x) | xxxx (xx.x) | xxxx (xx.x) | xxxx (xx.x) | | | xxxx (xx.x) | xxxx (xx.x) | | |
| Multi-domain | xxxx (xx.x) | xxxx (xx.x) | xxxx (xx.x) | xxxx (xx.x) | | | xxxx (xx.x) | xxxx (xx.x) | | |
| Mild | xxxx (xx.x) | xxxx (xx.x) | xxxx (xx.x) | xxxx (xx.x) | | | xxxx (xx.x) | xxxx (xx.x) | | |
| Moderate | xxxx (xx.x) | xxxx (xx.x) | xxxx (xx.x) | xxxx (xx.x) | | | xxxx (xx.x) | xxxx (xx.x) | | |
| Severe | xxxx (xx.x) | xxxx (xx.x) | xxxx (xx.x) | xxxx (xx.x) | | | xxxx (xx.x) | xxxx (xx.x) | | |

| | | | | | | | | |
|---|---|---|---|---|---|---|---|---|
| Death | xxxx (xx.x) | xxxx (xx.x) | xxxx (xx.x) | xxxx (xx.x) | | xxxx (xx.x) | xxxx (xx.x) | |
| **Primary, complete data** | | | | | | | | |
| Cognition - 7 level, N=1178 | xx.x [xx.x,xx.x] | xx.x [xx.x,xx.x] | xx.x [xx.x,xx.x] | xx.x [xx.x,xx.x] | MWUA | xx.x [xx.x,xx.x] | xx.x [xx.x,xx.x] | aOLR |
| Normal | xxxx (xx.x) | xxxx (xx.x) | xxxx (xx.x) | xxxx (xx.x) | | xxxx (xx.x) | xxxx (xx.x) | |
| Single-domain | xxxx (xx.x) | xxxx (xx.x) | xxxx (xx.x) | xxxx (xx.x) | | xxxx (xx.x) | xxxx (xx.x) | |
| Multi-domain | xxxx (xx.x) | xxxx (xx.x) | xxxx (xx.x) | xxxx (xx.x) | | xxxx (xx.x) | xxxx (xx.x) | |
| Mild | xxxx (xx.x) | xxxx (xx.x) | xxxx (xx.x) | xxxx (xx.x) | | xxxx (xx.x) | xxxx (xx.x) | |
| Moderate | xxxx (xx.x) | xxxx (xx.x) | xxxx (xx.x) | xxxx (xx.x) | | xxxx (xx.x) | xxxx (xx.x) | |
| Severe | xxxx (xx.x) | xxxx (xx.x) | xxxx (xx.x) | xxxx (xx.x) | | xxxx (xx.x) | xxxx (xx.x) | |
| Death | xxxx (xx.x) | xxxx (xx.x) | xxxx (xx.x) | xxxx (xx.x) | | xxxx (xx.x) | xxxx (xx.x) | |
| **Secondary, all data** | | | | | | | | |
| Cognition - 4 level | xx.x [xx.x,xx.x] | xx.x [xx.x,xx.x] | xx.x [xx.x,xx.x] | xx.x [xx.x,xx.x] | MWUA | xx.x [xx.x,xx.x] | xx.x [xx.x,xx.x] | aOLR |
| Normal | xxxx (xx.x) | xxxx (xx.x) | xxxx (xx.x) | xxxx (xx.x) | | xxxx (xx.x) | xxxx (xx.x) | |
| Minor | xxxx (xx.x) | xxxx (xx.x) | xxxx (xx.x) | xxxx (xx.x) | | xxxx (xx.x) | xxxx (xx.x) | |
| Dementia | xxxx (xx.x) | xxxx (xx.x) | xxxx (xx.x) | xxxx (xx.x) | | xxxx (xx.x) | xxxx (xx.x) | |
| Death | xxxx (xx.x) | xxxx (xx.x) | xxxx (xx.x) | xxxx (xx.x) | | xxxx (xx.x) | xxxx (xx.x) | |

| | | | | | | | | |
|---|---|---|---|---|---|---|---|---|
| Memory or thinking problem (%) | xxxx (xx.x) | xxxx (xx.x) | xxxx (xx.x) | xxxx (xx.x) | MWUA | xxxx (xx.x) | xxxx (xx.x) | aBLR |
| MoCA-visit | xx.x (xx.x) | xx.x (xx.x) | - | - | - | | | |
| MoCA-t | xx.x (xx.x) | xx.x (xx.x) | xx.x (xx.x) | xx.x (xx.x) | MWUA | xx.x (xx.x) | xx.x (xx.x) | aMLR |
| TICS-M | xx.x (xx.x) | xx.x (xx.x) | xx.x (xx.x) | xx.x (xx.x) | MWUA | xx.x (xx.x) | xx.x (xx.x) | aMLR |
| Trails A, time | xx.x (xx.x) | xx.x (xx.x) | - | - | MWU | - | - | - |
| Trails A, points | xx.x (xx.x) | xx.x (xx.x) | - | - | MWU | - | - | - |
| Trails B, time | xx.x (xx.x) | xx.x (xx.x) | - | - | MWU | - | - | - |
| Trails B, points | xx.x (xx.x) | xx.x (xx.x) | - | - | MWU | - | - | - |
| Verbal fluency phonemic (F) | xx.x (xx.x) | xx.x (xx.x) | xx.x (xx.x) | xx.x (xx.x) | MWUA | xx.x (xx.x) | xx.x (xx.x) | aMLR |
| Verbal fluency phonemic (letter A) | - | xx.x (xx.x) | xx.x (xx.x) | xx.x (xx.x) | MWUA | - | - | - |
| Verbal fluency phonemic (letter S) | - | xx.x (xx.x) | xx.x (xx.x) | xx.x (xx.x) | MWUA | - | - | - |
| Verbal fluency (Animal naming) | - | xx.x (xx.x) | xx.x (xx.x) | xx.x (xx.x) | MWUA | - | - | - |
| ***Cognitive domains*** | | | | | | | | |
| **Complex attention** | xx.x (xx.x) | xx.x (xx.x) | xx.x (xx.x) | xx.x (xx.x) | MWUA | xx.x (xx.x) | xx.x (xx.x) | aMLR |
| **Executive function** | xx.x (xx.x) | xx.x (xx.x) | xx.x (xx.x) | xx.x (xx.x) | MWUA | xx.x (xx.x) | xx.x (xx.x) | aMLR |
| **Learning & memory** | xx.x (xx.x) | xx.x (xx.x) | xx.x (xx.x) | xx.x (xx.x) | MWUA | xx.x (xx.x) | xx.x (xx.x) | aMLR |
| **Language** | xx.x (xx.x) | xx.x (xx.x) | xx.x (xx.x) | xx.x (xx.x) | MWUA | xx.x (xx.x) | xx.x (xx.x) | aMLR |
| **Perceptual-motor function** | xx.x (xx.x) | xx.x (xx.x) | xx.x (xx.x) | xx.x (xx.x) | MWUA | xx.x (xx.x) | xx.x (xx.x) | aMLR |
| **Social cognition** | xx.x (xx.x) | xx.x (xx.x) | xx.x (xx.x) | xx.x (xx.x) | MWUA | xx.x (xx.x) | xx.x (xx.x) | aMLR |

| | | | | | | | | |
|---|---|---|---|---|---|---|---|---|
| Dementia, clinical diagnosis | xxxx (xx.x) | xxxx (xx.x) | xxxx (xx.x) | xxxx (xx.x) | aBRM | xxxx (xx.x) | xxxx (xx.x) | aBLR |
| **Cognition by informant** | | | | | | | | |
| IQCODE, N=286 | xx.x (xx.x) | xx.x (xx.x) | xx.x (xx.x) | xx.x (xx.x) | MWUA | xx.x (xx.x) | xx.x (xx.x) | aMLR |
| 4AT | xxxx (xx.x) | xxxx (xx.x) | xxxx (xx.x) | xxxx (xx.x) | MWUA | xxxx (xx.x) | xxxx (xx.x) | aBLR |
| Apathy | xxxx (xx.x) | xxxx (xx.x) | xxxx (xx.x) | xxxx (xx.x) | MWUA | xxxx (xx.x) | xxxx (xx.x) | aBLR |
| Zarit score | - | - | xx.x (xx.x) | xx.x (xx.x) | MWU | xx.x (xx.x) | xx.x (xx.x) | aMLR |
| modified Rankin Scale (mRS) | xx.x [xx.x,xx.x] | xx.x [xx.x,xx.x] | xx.x [xx.x,xx.x] | xx.x [xx.x,xx.x] | MWUA | xx.x [xx.x,xx.x] | xx.x [xx.x,xx.x] | aOLR |
| Barthel index (BI) | xx.x (xx.x) | xx.x (xx.x) | xx.x (xx.x) | xx.x (xx.x) | MWUA | xx.x (xx.x) | xx.x (xx.x) | aMLR |
| Lawton ADL | xx.x (xx.x) | xx.x (xx.x) | xx.x (xx.x) | xx.x (xx.x) | MWUA | xx.x (xx.x) | xx.x (xx.x) | aMLR |
| Clinical frailty score (CFS) | xx.x (xx.x) | xx.x (xx.x) | xx.x (xx.x) | xx.x (xx.x) | MWUA | xx.x (xx.x) | xx.x (xx.x) | aOLR |
| **Activities of daily living** | | | | | | | | |
| Barthel index (BI) | xx.x (xx.x) | xx.x (xx.x) | xx.x (xx.x) | xx.x (xx.x) | MWUA | xx.x (xx.x) | xx.x (xx.x) | aMLR |
| Lawton ADL | xx.x (xx.x) | xx.x (xx.x) | xx.x (xx.x) | xx.x (xx.x) | MWUA | xx.x (xx.x) | xx.x (xx.x) | aMLR |
| Brief physical activity assessment, vigorous | xx.x (xx.x) | - | xx.x (xx.x) | xx.x (xx.x) | MWUA | xx.x (xx.x) | xx.x (xx.x) | aOLR |
| Brief physical activity assessment, moderate | xx.x (xx.x) | - | xx.x (xx.x) | xx.x (xx.x) | MWUA | xx.x (xx.x) | xx.x (xx.x) | aOLR |
| Short stroke impact scale | | | | | | | | |

| | | | | | | | | |
|---|---|---|---|---|---|---|---|---|
| Strength | - | xx.x (xx.x) | xx.x (xx.x) | xx.x (xx.x) | MWUA | xx.x (xx.x) | xx.x (xx.x) | aOLR |
| Memory | - | xx.x (xx.x) | xx.x (xx.x) | xx.x (xx.x) | MWUA | xx.x (xx.x) | xx.x (xx.x) | aOLR |
| Emotion | - | xx.x (xx.x) | xx.x (xx.x) | xx.x (xx.x) | MWUA | xx.x (xx.x) | xx.x (xx.x) | aOLR |
| Communication | - | xx.x (xx.x) | xx.x (xx.x) | xx.x (xx.x) | MWUA | xx.x (xx.x) | xx.x (xx.x) | aOLR |
| ADL | - | xx.x (xx.x) | xx.x (xx.x) | xx.x (xx.x) | MWUA | xx.x (xx.x) | xx.x (xx.x) | aOLR |
| Mobility | - | xx.x (xx.x) | xx.x (xx.x) | xx.x (xx.x) | MWUA | xx.x (xx.x) | xx.x (xx.x) | aOLR |
| Hand | - | xx.x (xx.x) | xx.x (xx.x) | xx.x (xx.x) | MWUA | xx.x (xx.x) | xx.x (xx.x) | aOLR |
| Social | - | xx.x (xx.x) | xx.x (xx.x) | xx.x (xx.x) | MWUA | xx.x (xx.x) | xx.x (xx.x) | aOLR |
| Global | - | - | - | - | | - | - | WLT |
| **Fatigue** | | | | | | | | |
| Brief fatigue inventory (BFI) | xxxx (xx.x) | xxxx (xx.x) | xxxx (xx.x) | xxxx (xx.x) | MWUA | xxxx (xx.x) | xxxx (xx.x) | aBLR |
| **Frailty** | | | | | | | | |
| Clinical frailty score (CFS) | xx.x [xx.x,xx.x] | xx.x [xx.x,xx.x] | xx.x [xx.x,xx.x] | xx.x [xx.x,xx.x] | MWUA | xx.x [xx.x,xx.x] | xx.x [xx.x,xx.x] | aMLR |
| **Functional** | | | | | | | | |
| modified Rankin Scale (mRS) | xx.x [xx.x,xx.x] | xx.x [xx.x,xx.x] | xx.x [xx.x,xx.x] | xx.x [xx.x,xx.x] | MWUA | xx.x [xx.x,xx.x] | xx.x [xx.x,xx.x] | aOLR |
| Disposition | xx.x [xx.x,xx.x] | xx.x [xx.x,xx.x] | xx.x [xx.x,xx.x] | xx.x [xx.x,xx.x] | MWUA | xx.x [xx.x,xx.x] | xx.x [xx.x,xx.x] | aOLR |
| **Mood (apathy, anxiety, depression)** | | | | | | | | |
| PHQ-2 | xx.x [xx.x,xx.x] | xx.x [xx.x,xx.x] | xx.x [xx.x,xx.x] | xx.x [xx.x,xx.x] | MWUA | xx.x [xx.x,xx.x] | xx.x [xx.x,xx.x] | aOLR |
| PHQ-9 | xx.x (xx.x) | xx.x (xx.x) | xx.x (xx.x) | xx.x (xx.x) | MWUA | xx.x (xx.x) | xx.x (xx.x) | aMLR |
| Generalised anxiety disorder (GAD) | xx.x (xx.x) | xx.x (xx.x) | xx.x (xx.x) | xx.x (xx.x) | MWUA | xx.x (xx.x) | xx.x (xx.x) | aMLR |
| Zung depression scale (ZDS) | xx.x (xx.x) | xx.x (xx.x) | xx.x (xx.x) | xx.x (xx.x) | MWUA | xx.x (xx.x) | xx.x (xx.x) | aMLR |

| | | | | | | | | |
|---|---|---|---|---|---|---|---|---|
| Office National Statistics-4 (ONS-4) | - | xx.x (xx.x) | xx.x (xx.x) | xx.x (xx.x) | MWUA | xx.x (xx.x) | xx.x (xx.x) | aMLR |
| Depression, clinical diagnosis | xxxx (xx.x) | xxxx (xx.x) | xxxx (xx.x) | xxxx (xx.x) | BRM | xxxx (xx.x) | xxxx (xx.x) | aBLR |
| **Quality of life** | | | | | | | | |
| EQ-5D-5L, as HU | xx.x (xx.x) | xx.x (xx.x) | xx.x (xx.x) | xx.x (xx.x) | MWUA | xx.x (xx.x) | xx.x (xx.x) | aMLR |
| EQ-VAS | - | xx.x (xx.x) | xx.x (xx.x) | xx.x (xx.x) | MWUA | xx.x (xx.x) | xx.x (xx.x) | aMLR |
| **Social support** | | | | | | | | |
| Social support survey (SSS) | xx.x (xx.x) | - | xx.x (xx.x) | xx.x (xx.x) | MWUA | xx.x (xx.x) | xx.x (xx.x) | aMLR |
| **Global** | | | | | | | | |
| Global overall | - | - | - | - | - | - | - | WLT |
| Global cognition | - | - | - | - | - | - | - | WLT |
| Global mood | - | - | - | - | - | - | - | WLT |
| Global mobility | - | - | - | - | - | - | - | WLT |
| Global neuro-psychiatric | - | - | - | - | - | - | - | WLT |
| **Vascular events (%)** | | | | | | | | |
| Recurrent stroke, TIA [7] | - | xxxx (xx.x) | xxxx (xx.x) | xxxx (xx.x) | BRM | xxxx (xx.x) | xxxx (xx.x) | aBLR |
| Myocardial infarction | - | xxxx (xx.x) | xxxx (xx.x) | xxxx (xx.x) | BRM | xxxx (xx.x) | xxxx (xx.x) | aBLR |
| **Risk factor status (%)** | | | | | | | | |
| Atrial fibrillation (AF) | xxxx (xx.x) | xxxx (xx.x) | xxxx (xx.x) | xxxx (xx.x) | BRM | xxxx (xx.x) | xxxx (xx.x) | aBLR |
| Diabetes mellitus (DM) | xxxx (xx.x) | xxxx (xx.x) | xxxx (xx.x) | xxxx (xx.x) | BRM | xxxx (xx.x) | xxxx (xx.x) | aBLR |

| | | | | | | | | |
|---|---|---|---|---|---|---|---|---|
| High blood pressure (HT) | xxxx (xx.x) | - | xxxx (xx.x) | xxxx (xx.x) | BRM | xxxx (xx.x) | xxxx (xx.x) | aBLR |
| Blood pressure (mmHg) | | | | | | | | |
| Systolic | xxx.x (xx.x) | xxx.x (xx.x) | - | - | MWU | - | - | - |
| Diastolic | xx.x (xx.x) | xx.x (xx.x) | - | - | MWU | - | - | - |
| Heart rate (bpm) | xxxx (xx.x) | xxxx (xx.x) | - | - | MWU | - | - | - |
| **Lifestyle** | | | | | | | | |
| Smoking, current | xxxx (xx.x) | xxxx (xx.x) | xxxx (xx.x) | xxxx (xx.x) | BRM | xxxx (xx.x) | xxxx (xx.x) | aBLR |
| Alcohol, >21 upw | xxxx (xx.x) | xxxx (xx.x) | xxxx (xx.x) | xxxx (xx.x) | BRM | xxxx (xx.x) | xxxx (xx.x) | aBLR |
| Salt, added | xxxx (xx.x) | xxxx (xx.x) | xxxx (xx.x) | xxxx (xx.x) | BRM | xxxx (xx.x) | xxxx (xx.x) | aBLR |

MoCA-m: Montreal cognitive assessment-modified; MoCA-t: Montreal cognitive assessment-telephone; PHQ: Patient health questionnaire; TICS-M: Telephone interview cognitive status

**Table 2. Pairwise comparisons of DSM5 at 0 versus 6, 6 versus 52 and 52 versus 104 weeks**

Data are median [interquartile range]. Comparisons by Mann-Whitney U test (all data) and Wilcoxon paired test (complete data).

| Timepoints (weeks) | Timepoint 1 | Timepoint 2 | Difference (95% CI) | 2p |
|---|---|---|---|---|
| **All data** | | | | |
| 0 vs 6 | xx.x [xx.x,xx.x] | xx.x [xx.x,xx.x] | xx.x (xx.x, xx.x) | MWU |
| 6 vs 52 | xx.x [xx.x,xx.x] | xx.x [xx.x,xx.x] | xx.x (xx.x, xx.x) | MWU |
| 52 vs 104 | xx.x [xx.x,xx.x] | xx.x [xx.x,xx.x] | xx.x (xx.x, xx.x) | MWU |
| **Complete data** | | | | |
| 0 vs 6 | xx.x [xx.x,xx.x] | xx.x [xx.x,xx.x] | xx.x (xx.x, xx.x) | WPT |
| 6 vs 52 | xx.x [xx.x,xx.x] | xx.x [xx.x,xx.x] | xx.x (xx.x, xx.x) | WPT |
| 52 vs 104 | xx.x [xx.x,xx.x] | xx.x [xx.x,xx.x] | xx.x (xx.x, xx.x) | WPT |

**Table 3. Prediction of DSM5-7 level cognition at two years from baseline characteristics**. Data are odds ratio (95% confidence intervals) using ordinal logistic regression: (A) Univariate; (B) Multiple variable; (C) Multiple variable, stepdown analysis; with adjustment for all listed covariates.

| | Univariate OLR | p | Multiple variable OLR-M coefficients | p | Stepdown OLR-M coefficients | p |
|---|---|---|---|---|---|---|
| Demographic | | | | | | |
| Age | x.xxx | x | x.xxx | x | x.xxx | x |
| Sex | x.xxx | x | x.xxx | x | x.xxx | x |
| Ethnicity, non-white | x.xxx | x | x.xxx | x | x.xxx | x |
| Highest education /6 | x.xxx | x | x.xxx | x | x.xxx | x |
| Marital status | x.xxx | x | x.xxx | x | x.xxx | x |
| Employment /3 | x.xxx | x | x.xxx | x | x.xxx | x |
| Social support /20 | x.xxx | x | x.xxx | x | x.xxx | x |
| Pre-morbid mRS /5 | x.xxx | x | x.xxx | x | x.xxx | x |
| Capacity | x.xxx | x | x.xxx | x | x.xxx | x |
| History | | | | | | |
| Alcohol | x.xxx | x | x.xxx | x | x.xxx | x |
| History of hypertension | x.xxx | x | x.xxx | x | x.xxx | x |
| History of stroke/TIA | x.xxx | x | x.xxx | x | x.xxx | x |
| Atrial fibrillation | x.xxx | x | x.xxx | x | x.xxx | x |
| Stroke related | | | | | | |
| Index stroke (ICH/IS/TIA) | x.xxx | x | x.xxx | x | x.xxx | x |
| Side of signs | x.xxx | x | x.xxx | x | x.xxx | x |
| NIHSS | x.xxx | x | x.xxx | x | x.xxx | x |
| Systolic blood pressure | x.xxx | x | x.xxx | x | x.xxx | x |
| Syndrome (cortical/lacunar/posterior) | x.xxx | x | x.xxx | x | x.xxx | x |
| Montreal Cognitive Assessment (MoCA) | x.xxx | x | x.xxx | x | x.xxx | x |
| Imaging | | | | | | |
| Visible acute lesion | x.xxx | x | x.xxx | x | x.xxx | x |
| Brain frailty score | x.xxx | x | x.xxx | x | x.xxx | x |
| SVD score | x.xxx | x | x.xxx | x | x.xxx | x |

**Table 4. Comparison of outcomes at two years between index events.**
Data are number (%), median [interquartile range] or mean (standard deviation). Comparisons by stroke type calculated using Chi-square test (CST), Kruskal-Wallis test (KWT) or one-way analysis of variance (ANOVA). Global analyses were performed using the Wei-Lachin test (WLT).

| **Week**<br>Number | **ICH** | **IS** | **TIA** | **p** |
|---|---|---|---|---|
| **Cognition, participant** | | | | |
| **Primary** | | | | |
| Cognition - 7 level | | | | KWT |
| Normal | xxxx (xx.x) | xxxx (xx.x) | xxxx (xx.x) | |
| Single-domain | xxxx (xx.x) | xxxx (xx.x) | xxxx (xx.x) | |
| Multi-domain | xxxx (xx.x) | xxxx (xx.x) | xxxx (xx.x) | |
| Mild | xxxx (xx.x) | xxxx (xx.x) | xxxx (xx.x) | |
| Moderate | xxxx (xx.x) | xxxx (xx.x) | xxxx (xx.x) | |
| Severe | xxxx (xx.x) | xxxx (xx.x) | xxxx (xx.x) | |
| Death | xxxx (xx.x) | xxxx (xx.x) | xxxx (xx.x) | CST |
| **Secondary** | | | | |
| Cognition - 4 level | | | | KWT |
| Normal | xxxx (xx.x) | xxxx (xx.x) | xxxx (xx.x) | |
| Minor | xxxx (xx.x) | xxxx (xx.x) | xxxx (xx.x) | |
| Dementia | xxxx (xx.x) | xxxx (xx.x) | xxxx (xx.x) | |
| Death | xxxx (xx.x) | xxxx (xx.x) | xxxx (xx.x) | - |
| Memory or thinking problem | xxxx (xx.x) | xxxx (xx.x) | xxxx (xx.x) | CST |
| Telephone-Montreal cognitive assessment (T-MoCA) | xx.x (xx.x) | xx.x (xx.x) | xx.x (xx.x) | ANOVA |
| Telephone interview cognitive status (TICS-M) | xx.x (xx.x) | xx.x (xx.x) | xx.x (xx.x) | ANOVA |
| Verbal fluency phonemic (letter F) | xx.x (xx.x) | xx.x (xx.x) | xx.x (xx.x) | ANOVA |
| Verbal fluency phonemic (letter A) | xx.x (xx.x) | xx.x (xx.x) | xx.x (xx.x) | ANOVA |
| Verbal fluency phonemic (letter S) | xx.x (xx.x) | xx.x (xx.x) | xx.x (xx.x) | ANOVA |
| Verbal fluency (Animal naming) | xx.x (xx.x) | xx.x (xx.x) | xx.x (xx.x) | ANOVA |
| Dementia, clinical diagnosis | xxxx (xx.x) | xxxx (xx.x) | xxxx (xx.x) | CST |
| **Informant** | | | | |
| IQCODE | xx.x (xx.x) | xx.x (xx.x) | xx.x (xx.x) | ANOVA |

| | | | | |
|---|---|---|---|---|
| 4AT | xxxx (xx.x) | xxxx (xx.x) | xxxx (xx.x) | KWT |
| NPI-Q | xxxx (xx.x) | xxxx (xx.x) | xxxx (xx.x) | KWT |
| Apathy | xxxx (xx.x) | xxxx (xx.x) | xxxx (xx.x) | KWT |
| Zarit score | xx.x (xx.x) | xx.x (xx.x) | xx.x (xx.x) | ANOVA |
| modified Rankin Scale (mRS) | xx.x (xx.x) | xx.x (xx.x) | xx.x (xx.x) | KWT |
| Barthel index (BI) | xx.x (xx.x) | xx.x (xx.x) | xx.x (xx.x) | ANOVA |
| Clinical frailty score (CFS) | xx.x (xx.x) | xx.x (xx.x) | xx.x (xx.x) | KWT |
| Lawton ADL | xx.x (xx.x) | xx.x (xx.x) | xx.x (xx.x) | ANOVA |
| **Activities of daily living** | | | | |
| Barthel index (BI) | xx.x (xx.x) | xx.x (xx.x) | xx.x (xx.x) | ANOVA |
| Lawton ADL | xx.x (xx.x) | xx.x (xx.x) | xx.x (xx.x) | ANOVA |
| Brief physical activity assessment, Vigorous | xx.x (xx.x) | xx.x (xx.x) | xx.x (xx.x) | KWT |
| Brief physical activity assessment, Moderate | xx.x (xx.x) | xx.x (xx.x) | xx.x (xx.x) | KWT |
| Short form SIS - Strength | xx.x (xx.x) | xx.x (xx.x) | xx.x (xx.x) | KWT |
| Short form SIS - Memory | xx.x (xx.x) | xx.x (xx.x) | xx.x (xx.x) | KWT |
| Short form SIS - Emotion | xx.x (xx.x) | xx.x (xx.x) | xx.x (xx.x) | KWT |
| Short form SIS - Communication | xx.x (xx.x) | xx.x (xx.x) | xx.x (xx.x) | KWT |
| Short form SIS - ADL | xx.x (xx.x) | xx.x (xx.x) | xx.x (xx.x) | KWT |
| Short form SIS - Mobility | xx.x (xx.x) | xx.x (xx.x) | xx.x (xx.x) | KWT |
| Short form SIS - Hand | xx.x (xx.x) | xx.x (xx.x) | xx.x (xx.x) | KWT |
| Short form SIS - Social | xx.x (xx.x) | xx.x (xx.x) | xx.x (xx.x) | KWT |
| **Fatigue** | | | | |
| Brief fatigue inventory (BFI) | xxxx (xx.x) | xxxx (xx.x) | xxxx (xx.x) | CST |
| **Frailty** | | | | |
| Clinical frailty score (CFS) | xx.x (xx.x) | xx.x (xx.x) | xx.x (xx.x) | KWT |
| **Functional** | | | | |
| modified Rankin Scale (mRS) | xx.x (xx.x) | xx.x (xx.x) | xx.x (xx.x) | KWT |
| Disposition | xx.x (xx.x) | xx.x (xx.x) | xx.x (xx.x) | KWT |
| **Mood (apathy, anxiety, depression)** | | | | |

| | | | | |
|---|---|---|---|---|
| Patient health questionnaire-2 (PHQ-2) | xx.x (xx.x) | xx.x (xx.x) | xx.x (xx.x) | ANOVA |
| Patient health questionnaire-9 (PHQ-9) | xx.x (xx.x) | xx.x (xx.x) | xx.x (xx.x) | ANOVA |
| Generalised anxiety disorder (GAD) | xx.x (xx.x) | xx.x (xx.x) | xx.x (xx.x) | ANOVA |
| Zung depression scale (ZDS) | xx.x (xx.x) | xx.x (xx.x) | xx.x (xx.x) | ANOVA |
| Office National Statistics-4 (ONS-4) | xx.x (xx.x) | xx.x (xx.x) | xx.x (xx.x) | ANOVA |
| Depression, clinical diagnosis | xxxx (xx.x) | xxxx (xx.x) | xxxx (xx.x) | CST |
| **Quality of life** | | | | |
| EQ-5D-5L, as HU | xx.x (xx.x) | xx.x (xx.x) | xx.x (xx.x) | ANOVA |
| EQ-VAS | xx.x (xx.x) | xx.x (xx.x) | xx.x (xx.x) | ANOVA |
| **Social support** | | | | |
| Social support survey (SSS) | xx.x (xx.x) | xx.x (xx.x) | xx.x (xx.x) | ANOVA |
| **Global (ICH vs IS)** | | | | |
| Stroke impact scale (SIS) | | | | WLT |
| Global | | | | WLT |
| Global cognition | | | | WLT |
| Global mood | | | | WLT |
| Global mobility | | | | WLT |
| Global neuro-psychiatric | | | | WLT |
| **Vascular events (%)** | | | | |
| Recurrent stroke, TIA [7] | xxxx (xx.x) | xxxx (xx.x) | xxxx (xx.x) | CST |
| Myocardial infarction | xxxx (xx.x) | xxxx (xx.x) | xxxx (xx.x) | CST |
| **Risk factor status (%)** | | | | |
| Atrial fibrillation (AF) | xxxx (xx.x) | xxxx (xx.x) | xxxx (xx.x) | CST |
| Diabetes mellitus (DM) | xxxx (xx.x) | xxxx (xx.x) | xxxx (xx.x) | CST |
| High blood pressure (HT) | xxxx (xx.x) | xxxx (xx.x) | xxxx (xx.x) | CST |
| **Lifestyle** | | | | |
| Smoking, current | xxxx (xx.x) | xxxx (xx.x) | xxxx (xx.x) | CST |
| Alcohol, >21 upw | xxxx (xx.x) | xxxx (xx.x) | xxxx (xx.x) | CST |
| Salt, added | xxxx (xx.x) | xxxx (xx.x) | xxxx (xx.x) | CST |

**Figures**

1. 7- and 4-level ordinal cognition/dementia scale
2. STROBE flow diagram
3. Receiver operating characteristic (ROC) curves of 7-level cognition status versus adjudicated dementia
4. Trajectory of DSM5-7 level (box and whisker plot)
5. Trajectory of DSM5-4 level (box and whisker plot)
6. Horizontal stacked distributions of 7-level ordinal cognition by time points
7. Horizontal stacked distributions of mRS by time points
8. Box and whisker plot of cognition as MoCA at 2 years
9. Box and whisker plot of mood as PHQ-9 at 2 years
10. Box and whisker plot of anxiety as GAD at 2 years
11. Box and whisker plot of fatigue as BFI at 2 years
12. Box and whisker plot of apathy as NPI-Q at 2 years

**Figure 1**. Primary outcome cognition categories and operational definitions. Adapted from published R4VaD protocol.[1]

| Primary category =DSM V | Operationalisation | Sub-category | Operationalisation |
|---|---|---|---|
| **Normal cognition** | **No evidence of cognitive impairment**<br>(T-MoCA:20-22 AND TICS-m: 25-39) | Normal | |
| **Minor Neurocognitive disorder (mild cognitive impairment)** | **Evidence of cognitive impairment**<br>(T-MoCA: 15-19 OR TICS-m: 17-24)<br>**AND**<br>**No evidence of functional impairment**<br>(mRS <2 OR no change in mRS if pre-stroke mRS >1) | Single domain | Scores are reduced by > 1 point in only one cognitive domain of T-MoCA |
| | | Multi domain | Scores are reduced by > 1 point in more than one cognitive domain of T-MoCA |
| **Major neurocognitive disorder** | Evidence of persisting multi-domain cognitive impairment<br>(T-MoCA score <19 OR TICS-m<24 on more than one follow-up)<br>and<br>Evidence of functional impairment<br>(mRS ≥2 or IQCODE >3.6 at final follow-up)<br>(further categorised by severity in 7 level score)<br>OR<br>**Clinical diagnosis made independent of study**<br>Any clinical diagnosis of dementia made by memory clinic<br>(or equivalent, this would include primary care)<br><br>Any recording of dementia on death certification<br><br>Any prescription of cholinesterase inhibitor or memantine<br><br>**OR**<br>**Pre-stroke dementia**<br>(Baseline assessment IQCODE >3.6 AND MoCA <23) | Mild | **Cognitive impairments**<br>(T-MoCA 15-19 OR TICS-m 17-23)<br>**AND**<br>**Minimal functional problems**<br>(mRS <3) |
| | | Moderate | **More severe cognitive impairments**<br>(T-MoCA 10-14 OR TICS-m 12-16)<br>**AND**<br>**More limiting function**<br>(mRS 3 or 4) AND [Barthel >60 (if available)] |
| | | Severe | **Most severe cognitive impairments**<br>(T-MoCA <10 OR TICS-m <12)<br>**AND**<br>**Most limited function**<br>Care-home admission<br>OR<br>(mRS 4,5 OR Barthel <60)<br>OR<br>Any NPI Q item 3 |
| **Death** | | **Death** | |

'Equivalent' will include any formal diagnosis of dementia or a dementia subtype made by a suitably trained professional, in the UK this is likely to be a geriatrician, neurologist, old age psychiatrist or psychologist. At follow-up, the functional status (mRS and Barthel ADL) can be taken from either the participant or the informant, the more severe score should be used to inform the cognitive categorisation. MoCA can be used in place of T-MoCA where available.

**Figure 2. STROBE flow diagram**. Flow of patients through study at baseline, 6 weeks, 1 year and 2 years. Note: screening logs were not kept.

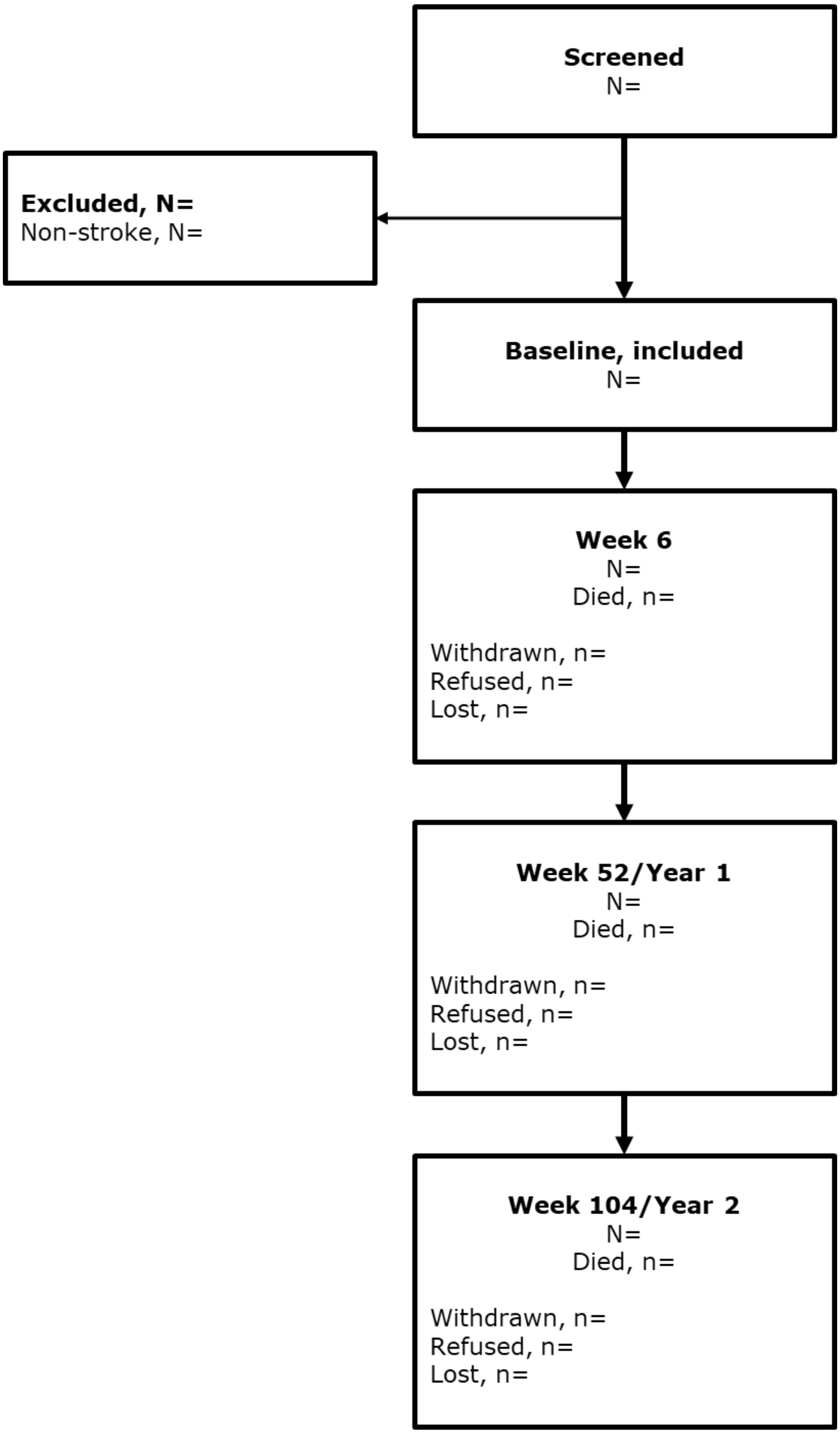

**Figure 3. Receiver operating characteristic (ROC) curves of 7-level cognition status versus adjudicated dementia**

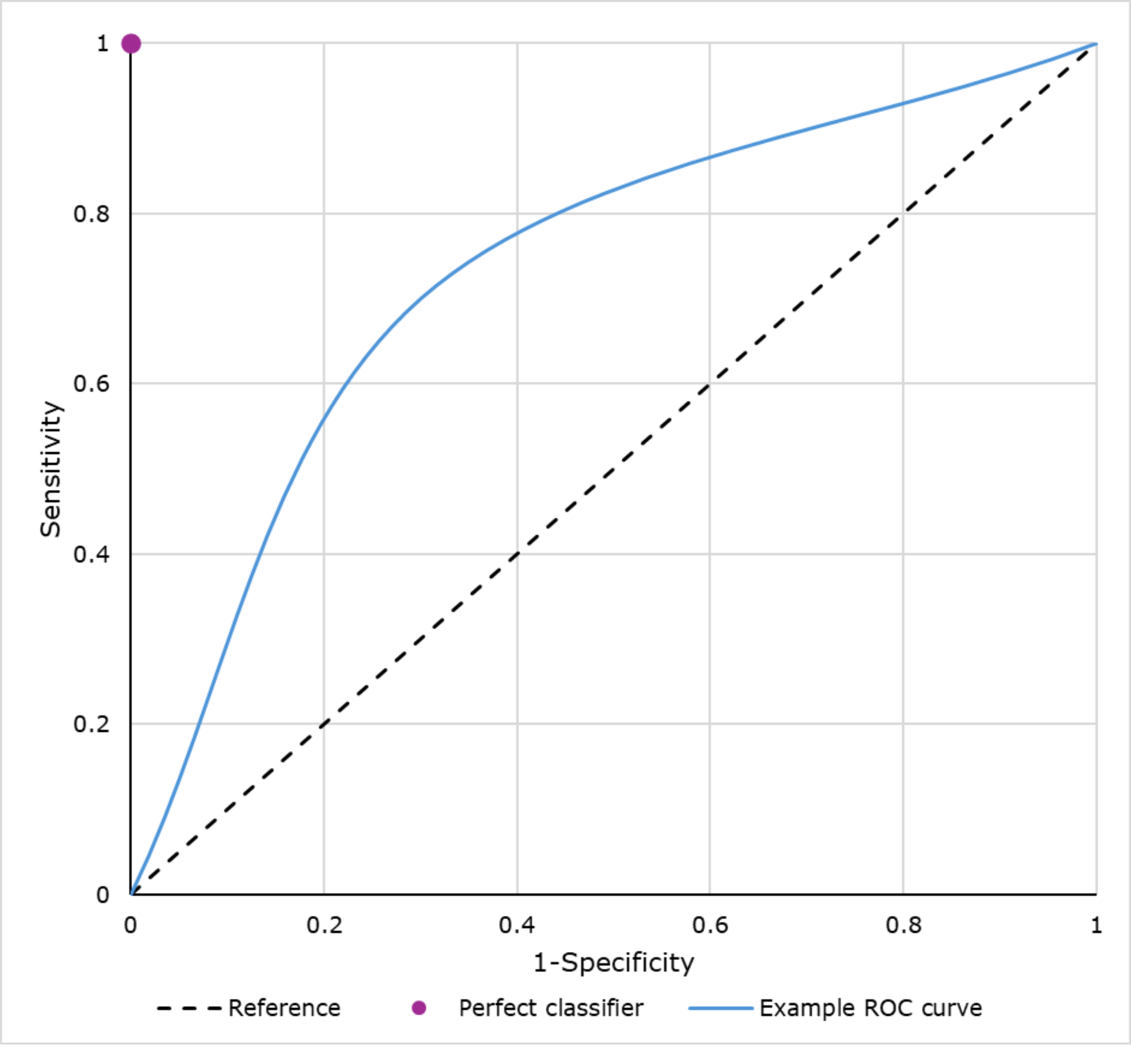
